# Chatbot as an intervention on physical activity: A systematic review and meta-analysis with meta-regression

**DOI:** 10.64898/2026.11.13.26358176

**Authors:** Lingyi Fu, Dongmin Song, Yang Bai

## Abstract

**Background:** Chatbots offer scalable, accessible, and personalized support for physical activity (PA). This review synthesized evidence on their efficacy and examined potential moderators.

**Methods:** Eleven databases were searched for English-language published through January 27, 2026. Eligibility was assessed using the PICOS framework. Data extraction was guided by research questions, mERA checklist, and PRISMA 2020. Risk of bias was evaluated using Cochrane Risk of Bias 2 tool and the NIH Quality Assessment Tool. Meta-analyses reported Hedges’ g. Publication bias was evaluated using funnel plots, Egger’s regression, and trim-and-fill analyses. Potential moderators were examined through subgroup analyses and meta-regression.

**Results:** A total of 12,481 records were identified, of which 43 studies were included in the systematic review and 27 in the meta-analysis. Most studies included in the meta-analysis had some risk-of-bias concerns (16/27). Chatbot interventions produced a significant small-to-moderate post-intervention effect on PA (g = 0.31, 95% CI [0.19, 0.44]; n = 24), whereas the follow-up effect was small and non-significant (g = 0.11, 95% CI [-0.01, 0.22]; n = 4). Effects varied across PA outcomes (g = 0.08–0.44), and several subgroup differences helped explain the significant heterogeneity. For daily steps, significant subgroup differences were observed by clinical status, while intervention duration (QM = 6.09, *p* = 0.048) and clinical status (QM = 5.69, *p* = 0.017) were significant moderators.

**Conclusions:** Chatbot interventions showed small-to-moderate post-intervention effects but limited follow-up effects. Few significant moderators were identified, underscoring the need for more rigorous RCTs.

## Introduction

Physical activity (PA) is widely recognized as critical components of public health [1,2] and has increasingly recognized as a leading modifiable exposure for controlling non-communicable diseases [3]. However, the prevalence of physical inactivity has risen significantly, from 23.4% in 2000 to 31.3% in 2022, with current trends indicating that the global target of a 15% reduction by 2030 is unlikely to be met [4]. Therefore, scalable strategies are needed to reduce physical inactivity at the national level. Digital health interventions, characterized by interventions delivered via digital technologies such as smartphones, websites, text messaging, have substantial potential to improve access to PA over the past decades [5]. In recent years, research has grown in interest in chatbots for promoting PA [6–8], especially with the advent of ChatGPT (Chat Generative Pre-Trained Transformer) in 2022. A Chatbot, otherwise known as “conversational agent”, is a computer program designed to respond to conversational or informational replies with verbal (audio-based chatbots) or written (text-based chatbots) messages [9]. These chatbots primarily functioned as health coaches or self-monitoring tools to promote PA [10].

Some existing meta-analytic evidence has demonstrated the moderate efficacy of chatbots for promoting PA [11,12]. However, some research gaps exist. First, limited synthesized evidence compares generative AI chatbots with traditional machine learning–based or rule-based chatbots for promoting PA. Computer-based health counseling emerged in the late 1990s [13,14], followed by early non-AI PA chatbots in the 2000s [15]. Since 2018, AI chatbots have become increasingly viable for PA support [10]. More recently, advances in generative AI since 2022 may further enhance chatbot-based PA interventions by reducing repetitive responses and increasing content novelty [16]. Therefore, updated evidence is needed to understand how advanced algorithms may influence chatbot performance in promoting PA. In addition to the evolution of chatbot algorithms, visual representations of chatbots, such as bodies and facial expressions, remain underexplored. These features may be important for enhancing user engagement during interactions. However, previous meta-analyses have not distinguished between chatbots with visual representations, such as embodied conversational agents, and chatbots without visual representations when examining intervention efficacy.

Regarding intervention characteristics, although intervention duration has been examined in previous meta-analyses [11,12], follow-up effects have not been explored. From public health and clinical perspectives, sustained effects at follow-up are important indicators of intervention efficacy. With the increasing number of chatbot intervention studies in recent years, the evidence base has expanded, allowing us to examine the difference between follow-up effects and post-intervention effects. Moreover, intervention efficacy may vary by comparison group. For example, previous studies suggested that non-visual AI chatbots may show different effects for promoting PA when compared with human coaches versus other chatbots with visual expression [10]. However, none of existing meta-analysis [11,12] explored how the efficacy varies from different control groups, such as usual care, human adviser, and base chatbot. Understanding how results differ by control condition is important because the comparison group directly shapes how intervention efficacy should be interpreted. Distinguishing between true controls, such as placebo or no treatment, and active controls helps determine whether chatbots are effective compared with doing nothing or offer added benefits over existing alternatives. In addition, context is also an important determinant of health outcomes [17]. However, to our knowledge, limited evidence is available on whether the efficacy of chatbots for promoting PA varies across different intervention settings such as community setting, free-living setting, and home setting. Lastly, from the participant characteristics perspective, previous reviews have highlighted the importance of examining whether chatbot efficacy differs between healthy and clinical populations [10], which may help inform the use of chatbots in clinical populations. However, there remains a lack of meta-analytic synthetic on the efficacy of chatbots among clinical status.

Therefore, this systematic review with meta-regression study aims to evaluate the efficacy of chatbot interventions on PA while also exploring potential moderators, including comparison group, multiple intervention, intervention duration, intervention setting, participants clinical status, assessment time point, chatbot output type, chatbot’s use of visual expression, chatbot’s algorithm type, as well as two groups of moderators including chatbot characteristics and intervention characteristics, through subgroup analyses and meta-regression.

## Methodology

### Protocol and registration

The protocol for this systematic review and meta-analysis was prospectively registered on Open Science Framework [18] and the results are reported according to PRISMA guidelines. There was no funding source for this study.

### Search strategy

We searched 11 databases, including Web of Science, PubMed, CINAHL, MEDLINE, EMBASE, Scopus, APA PsycINFO, IEEE Xplore, ACM Digital Library, arXiv, and medRxiv. The search strategy primarily focused on titles and abstracts, using terms related to chatbots and PA (see Table S1). To ensure a comprehensive search, we identified relevant synonyms of physical activity and chatbot from previously published studies. Filters were applied to include English-language records published before January 27, 2026. Document types were limited to articles, early access articles, and proceedings papers. One reviewer was involved in the data searching stages (L.F.).

### Selection process

The reference records were exported from each database in multiple formats (e.g., .ris, .nbib, and .enw) and then imported into *Covidence* platform for screening [19]. The study selection criteria were developed based on the PICOS framework: population (P; no restrictions), intervention (I; chatbot-based digital interventions), comparison (C; any comparison group), outcome (O; physical activity outcome only), and study design (S; randomized controlled trials, non-randomized controlled trials, and single-group pre–post studies). More details are provided in Table S2. We conducted two stages of screening: title/abstract screening and full-text screening. At each stage, two reviewers (L.F. and D.S.) independently screened the records. Disagreements were resolved through discussion and consultation from a third reviewer (Y.B.). Interrater reliability was calculated using Cohen’s kappa in Covidence [20]. The results indicated substantial agreement, with kappa values of 0.78 for title and abstract screening and 0.76 for full-text screening.

### Data extraction and synthesis

The original extraction items were derived based on the research questions, the mobile health evidence reporting and assessment (mERA) checklist [21], and the Preferred reporting items for systematic review and meta-analysis protocols (PRISMA-P) 2020 [22]. Extracted information included publication characteristics, chatbot characteristics, outcomes of interest, population characteristics, intervention characteristics, as well as sample sizes, means, and standard deviations data of intervention and control group (see Table S3 for more information). Included full-text articles were stored in Box folder, and data were extracted using a Microsoft Excel form. One reviewer (L.F.) conducted the primary extraction, while a second reviewer (D.S.) independently verified a random half of the articles. Discrepancies were resolved through discussion. A narrative approach was then used to systematically synthesize the evidence. All studies that met the review eligibility criteria were included in the narrative synthesis, and their study, outcome, participant, intervention, chatbot, characteristics were summarized in tables. Studies were included in the meta-analysis only when sufficient quantitative data were available to calculate an effect size.

### Risk of bias assessment

Two reviewers (L.F. and D.S.) independently assessed risk of bias. The review team was not blinded to the authors or journals during the review process. Disagreements were resolved through discussion between two reviewers. We used the Cochrane Risk of Bias 2 tool [23] to assess randomized controlled trials (RCTs), including parallel RCTs, cluster RCTs, and cross-over RCTs. The tool assesses five domains of bias: the randomization process, deviations from intended interventions, missing outcome data, outcome measurement, and selection of the reported result. Minor adaptations were applied for cluster RCTs and cross-over RCTs according to the Cochrane guidance. We followed the signaling questions and algorithms provided in the Cochrane documentation to determine the risk of bias for each domain and the overall risk of bias for each study. Overall risk of bias was classified as low risk, some concerns, or high risk. For single-group before–after studies, we used the NIH Quality Assessment Tool for pre–post studies with no control group [24,25]. This tool primarily assesses internal validity using 12 evaluation items, such as whether the study objective was clearly stated, eligibility criteria were prespecified, etc. For each study, we calculated the percentage of applicable items rated “Yes.” Consistent with previous studies [26], studies were classified as low risk (>75%), some risk (50%–75%), or high risk (<50%). To provide comprehensive findings, no studies were excluded based on risk-of-bias or quality ratings. The overall risk of bias for each study was finally visualized using the robvis tool [27]. Green indicates low risk, yellow indicates some concerns, and red indicates high risk.

### Outcomes of interest

Outcomes of interest were (1) daily steps; (2) moderate-to-vigorous physical activity (MVPA, minutes/week); (3) total physical activity minutes (any measure of low, moderate and/or vigorous physical activity reported as a duration), and other PA related outcomes if available.

### Meta-analysis methods

The main meta-analysis included both RCTs and single-group pre–post studies. An additional RCTs-only meta-analysis was conducted as a sensitive analysis. In both the main and sensitive analyses, two time points were analyzed separately: post-intervention and follow-up. Data were visualized by outcome type in the forest plots. Standardized mean differences (SMDs; Hedges’ g) were used as the effect measure to allow comparison across studies using different outcome scales. Hedges’ g was selected because it provides a bias-corrected estimate of effect size, particularly for studies with small sample sizes. Standardized classifications were used to interpret the magnitude of effect sizes: 0.20 as small, 0.20 to 0.50 as medium, and greater than 0.50 as large [28]. A p-value of < .05 was considered statistically significant. Statistical heterogeneity was assessed using the *I²* statistic. Tau-squared (τ²) was also reported to estimate between-study variance. The following cutoffs were used to interpret I²: 0% to 29% = no heterogeneity, 30% to 49% = moderate heterogeneity, 50% to 74% = substantial heterogeneity, and 75% to 100% = considerable heterogeneity [29]. All meta-analyses were conducted in R using the *metagen* function. We contacted study authors when sample size, means, and standard deviations were not reported in the article.

For publication bias analyses, effect sizes were aggregated within studies by outcome because multiple effect sizes derived from the same study are statistically dependent. This approach reduces potential violations of the independence assumption required for funnel plot interpretation, with each point in the funnel plot representing one outcome per study. Visual inspection of funnel plots and Egger’s regression test for funnel plot asymmetry was conducted. In addition, the trim-and-fill method was applied to estimate the potential impact of missing studies on the overall effect size. Publication bias analyses were conducted using the *metabias* function in R.

### Subgroup analysis and Meta-regression methods

Random-effects subgroup meta-analyses using restricted maximum likelihood were conducted separately for each PA outcome. Meta-regression was then used to identify differences between specific subgroup categories. Univariable and multivariable meta-regression analyses were conducted separately using restricted maximum likelihood. Multilevel random-effects meta-regression models were fitted to account for between-study heterogeneity and within-study dependence among effect sizes, as some studies contributed more than one comparison or effect size. For each moderator, we conducted outcome-specific and all-outcome models. Outcome-specific models were fitted separately when a sufficient number of studies were available for a given outcome (>10 studies) [30]. The all-outcome models pooled all PA-related outcomes to increase statistical power and provide a more comprehensive assessment of PA behavior. Analyses were performed in R using the *meta* package.

The examined moderators of subgroup meta-analysis and meta-regression included: (1) comparison group, including no intervention, usual care, human advisor, and text chatbot (base), (2) multiple intervention, categorized as chatbot only versus chatbot plus, (3) intervention duration, categorized as ≤6 weeks, >6 to <12 weeks, and ≥12 weeks, (4) intervention setting, including community, home, and free-living settings, (5) clinical status, categorized as clinical versus non-clinical, (6) assessment time point, categorized as mid-intervention, post-intervention, and follow-up, (7) chatbot output type, categorized as speech/voice, text, or mixed output, (8) use of visual expression, categorized as embodied conversational agent versus chatbot, and (9) algorithm type, categorized as non-AI, traditional AI, and generative AI. Categories within each moderator were included only if they were represented by more than one study. In addition, two groups of moderators were examined: chatbot characteristics, including output type, use of visual expression, and algorithm type; as well as intervention characteristics, including intervention duration, intervention setting, and clinical status.

## Results

### Literature search and screening

After the database search, 12,481 records were identified from 11 databases (see Figure 1). After removing 7,564 duplicates, 4,917 records were screened at the title and abstract stage. Of these, 232 full-text articles were assessed for eligibility. A total of 189 articles were excluded for the following reasons: wrong intervention (n = 54), wrong outcome (n = 70), wrong study design (n = 49), full text unavailable (n = 15), and wrong language (n =1). Finally, 43 studies were included in the systematic review. Among these, 27 studies were included in the meta-analysis. The remaining studies were excluded because they assessed PA-related psychosocial outcomes (n = 7), such as intention to change PA, PA self-efficacy, and PA habit strength, or did not provide sufficient data for meta-analysis (n = 9).

**Figure 1.**
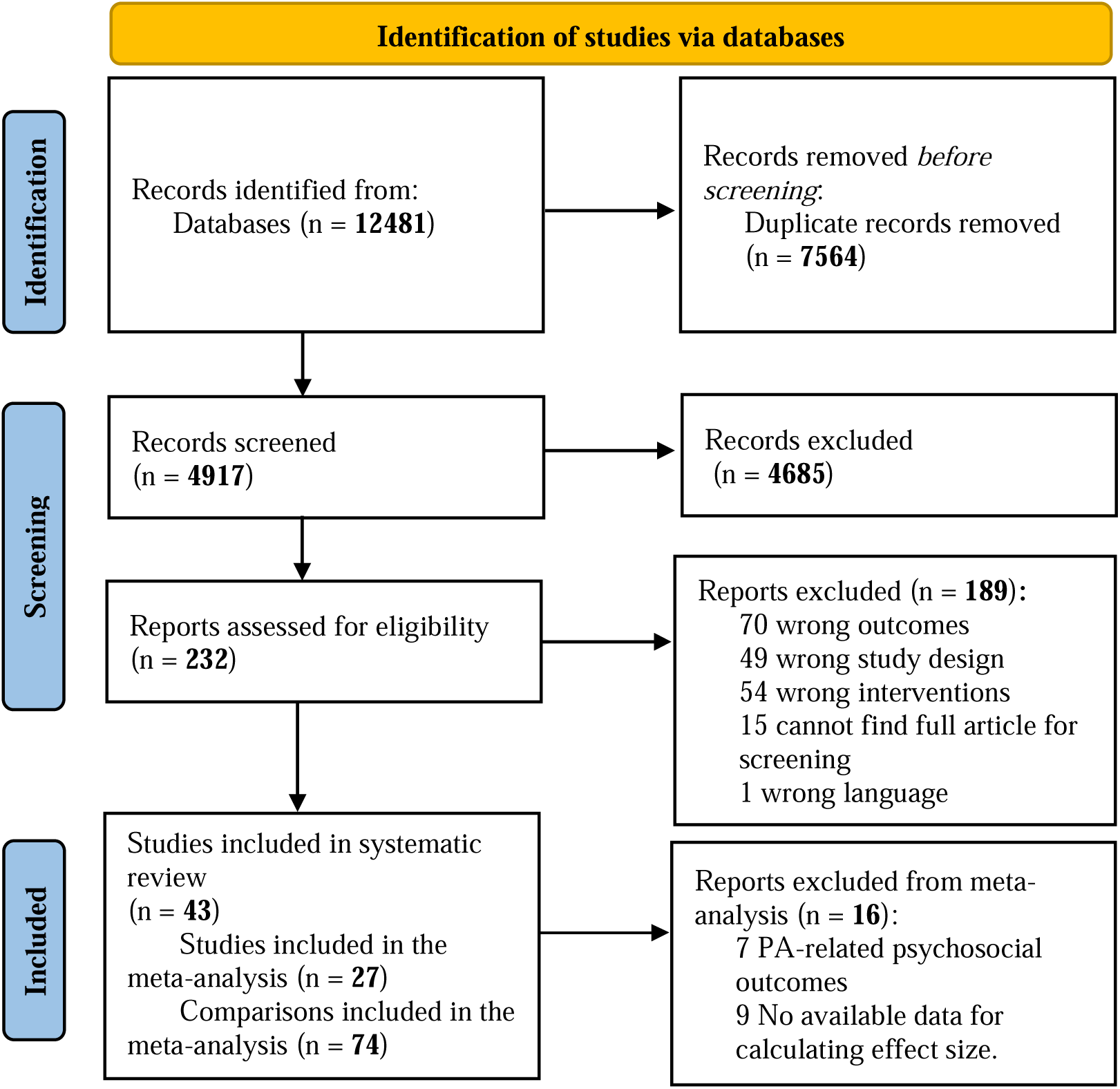
Literature selection process

### Study characteristics

Table S4 presents an overview of all included studies (n1 = 43) and the subset of studies included in the meta-analysis (n2 = 27). Table S5 provides more detailed information on outcomes and measurements, as well as descriptions of each study arm. In the following report, **n1** refers to the total number of studies included, whereas **n2** refers to the number of studies included in the meta-analysis.

The 43 studies included in the systematic review comprised 30 parallel RCTs (n2 = 17), one quasi-RCTs (n2 = 1/27), one cluster RCT (n2 = 1/27), one cross-over RCTs (n2 = 0/27), one micro-RCT (n2 = 0/27), and nine single-group studies (n2 = 8/27). An overview of the risk-of-bias assessment is provided in Figure S1 for RCTs and Figure S2 for single-group pre–post studies. Overall, most studies were judged as having some concerns (n1 = 25/43; n2 = 16/27), followed by high risk of bias (n = 12/43; n2 = 5/27), and low risk of bias (n1 = 6/43; n2 = 6/27). Among RCTs, the most common issues rated as “some concerns” were related to the randomization process, including insufficient reporting of randomization methods and unclear allocation concealment. Additional concerns were related to selective reporting, as many studies did not have a preregistered analysis plan. Substantial missing outcome data was the most common reason for studies being rated as high risk of bias. For single-group studies, common concerns included lack of outcome assessor blinding and limited pre–post measurement, which may reduce confidence in the reliability of the intervention effects.

Overall, the included studies were conducted across 12 countries or regions. The largest number of studies were conducted in the United States (n1 = 17/43; n2 = 12/27), followed by the Netherlands (n1 = 7/43; n2 = 4/27) and Australia (n1 = 5/43; n2 = 3/27). Publication years ranged from 2005 to 2026, with most studies published after 2020 (n1 = 31/43; n2 = 21/27). Dr. Timothy Bickmore’s research group contributed the largest number of studies on embodied conversational agents and PA (n1 = 9/43; n2 = 7/27). The total sample size was 5,901 across all included studies and 4,794 among studies included in the meta-analysis. Most studies recruited fewer than 100 participants (n1 = 25/43; n2 = 14/27). Participant age ranged from 12.02 to 74 years across all studies and from 12.02 to 71.3 years among studies included in the meta-analysis. Most studies focused on young and middle-aged adults aged 18 to 65 years (n1 = 33/43; n2 = 21/27). Female participants were overrepresented in many studies, with more than half of participants identifying as female in most samples (n1 = 33/43; n2 = 22/27). Regarding clinical status, most studies focused on non-clinical populations (n1 = 25/43; n2 = 17/27), while fewer targeted clinical or special populations (n1 = 10/43; n2 = 7/27). These included children with autism spectrum disorder [31], adults with overweight or obesity [32–34], cancer patients [35,36], individuals with chronic obstructive pulmonary disease [37] or hypertension [38], people with chronic conditions [39], and patients in clinical settings [40].

In terms of intervention characteristics, most studies combined chatbots with other intervention components (n1 = 28/43; n2 = 21/27), such as activity trackers, additional app functions, usual care, daily web-based educational content, home blood pressure monitoring, physician visits, or prescribed medications. Intervention duration ranged from a single session to 48 weeks. Most interventions lasted 12 weeks or less. Specifically, 19 studies lasting less than 6 weeks (n2 = 8/27), 18 studies lasting 6 to 12 weeks (n2 = 11/27), and 6 studies lasting more than 12 weeks (n2 = 6/27). Interaction frequency varied across studies and included daily, weekly, biweekly, several times per week, and several times per day. Among studies that specified a predefined interaction duration, each interaction typically lasted between 5 and 15 minutes. Most studies were conducted in free-living settings without a specified physical location (n1 = 24/43; n2 = 15/27). The remaining studies were conducted at home (n1 = 11/43; n2 = 8/27), in laboratories (n1 = 3/43; n2 = 1/27), community centers (n1 = 2/43; n2 = 2/27), schools (n1 = 1/43; n2 = 1/27), workplaces (n1 = 1/43; n2 = 0/27), and primary care clinics (n1 = 1/43; n2 = 0/27). Among studies in free-living settings (n1 = 24; n2 = 15), smartphones were the most used platform for interacting with the chatbot (n1 = 16/24; n2 = 11/15). Most smartphone-based interventions used standalone apps, although a small number integrated chatbots into existing social platforms, such as Slack [41,42] and Facebook Messenger [43]. In contrast, among studies conducted in specified physical settings (n1 = 19; n2 = 12), computers were the most commonly used platform (n1 = 15/19; n2 = 7/12).

Regarding chatbot characteristics, sixteen studies (n1 = 16/43) used chatbots that were graphically represented with a body or avatar and were categorized as embodied conversational agents (n2 = 8/27). Most embodied conversational agents used mixed text- and audio-based input/output modalities (n1 = 8/16; n2 = 6/8) and non-AI algorithms (n1 = 13/16; n2 = 8/8). Among non-embodied chatbots (n1 = 27/43; n2 = 19/27), most were text-based (n1 = 24/27; n2 = 16/19), two were audio-based, and one study compared text-based and audio-based chatbots [35]. Moreover, most non-embodied chatbots used traditional AI algorithms (n1 = 12/27; n2 = 10/19), followed by non-AI approaches (n1 = 7/27; n2 = 6/19) and generative AI approaches (n1 = 6/27; n2 = 3/19). One study combined generative AI, specifically large language modeling, with non-AI rule-based components [44]. Traditional AI-based chatbots commonly relied on commercial platforms, such as Amazon Alexa, IBM Watson, and Dialogflow. Whereas generative AI-based chatbots most often used GPT-series models with one study using Mistral [45].

In terms of measurement, steps were primarily assessed objectively among studies reporting step outcomes (n1 = 15/16; n2 = 13/14). One study used a pedometer to collect step counts but relied on participants to self-report the values [46]. In contrast, total PA was most often assessed using subjective measures (n1 = 9/11; n2 = 7/8), and MVPA was also primarily assessed subjectively (n1 = 8/9; n2 = 8/9). Other PA outcomes, including exercise duration, walking minutes, and PA goal achievement, were also mainly based on subjective reports.

### Meta-analyses and subgroup analysis results

Twenty-seven studies were included in the meta-analysis, of which three reported only follow-up effects.

#### Post-intervention effects

Figure 2 illustrates the post-intervention effects of chatbot interventions across different PA outcomes. Overall, chatbot interventions demonstrated a statistically significant medium effect on PA outcomes at post-intervention (g = 0.31, 95% CI [0.19, 0.44]; n = 24). Specifically, chatbot interventions showed a significant large effect on walking minutes (g = 1.24, 95% CI [0.99, 1.50]; n = 2), as well as significant medium effects on daily steps (g = 0.23, 95% CI [0.13, 0.32]; n = 11) and total PA (g = 0.44, 95% CI [0.06, 0.82]; n = 5). Non-significant effects were observed for exercise time (g = 0.34, 95% CI [-0.58, 1.25]; n = 4), MVPA (g = 0.08, 95% CI [-0.06, 0.21]; n = 7), and PA goal achievement (g = 0.02, 95% CI [-0.19, 0.22]; n = 1).

**Figure 2.**
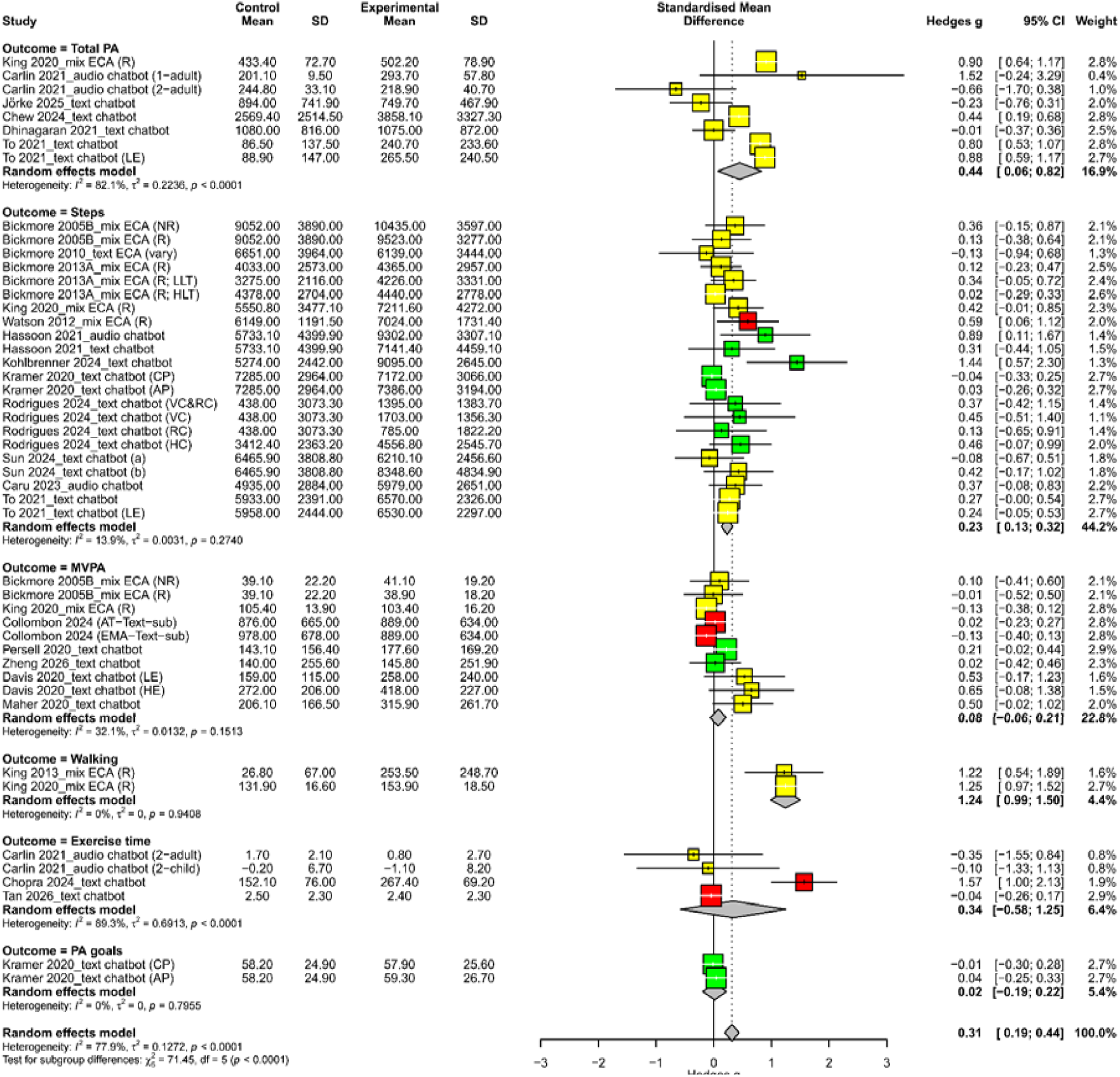
Post-intervention forest plot by outcome ***Note*.** Mix: chatbots that used both text- and audio-based communication; R: relational; NR: non-relational; 2-adult: phase 2 data for adults; 2-child: phase 2 data for children; LE: low engagement; HE: high engagement; Vary: dialogue structures that changed across conversations; LLT: low-literacy population; HLT: high-literacy population; CP: coping planning; AP: action planning; VC: visual cues; RC: relational cues; HC: human cues; a: non-humorous; b: humorous; AT: activity tracker; EMA: ecological momentary assessment; sub: subjective assessment.

Regarding heterogeneity in the post-intervention analyses, the significant heterogeneity was observed only for total PA (I² = 82.1%, *p* < 0.0001) and exercise minutes (I² = 89.3%, *p* < 0.0001). Subgroup analyses were further conducted to examine potential sources of heterogeneity, with results reported in Table S6. For total PA, significant subgroup differences were observed by visual expression (χ² = 4.71, df = 1, *p* = 0.030), algorithm type (χ² = 14.61, df = 2, *p* = 0.001), and comparison group (χ² = 13.93, df = 2, *p* = 0.001). For exercise minutes, significant subgroup differences were observed by multiple intervention (χ² = 27.88, df = 1, *p* = 0.000), comparison group (χ² = 11.73, df = 1, *p* = 0.001), and intervention setting (χ² = 11.73, df = 1, *p* = 0.001). These findings suggest that these factors may partly explain the large heterogeneity observed in total PA and exercise minutes. For outcomes with relatively small heterogeneity, significant subgroup differences were also observed. For steps, subgroup differences were statistically significant by clinical population status (χ² = 3.93, df = 1, *p* = 0.047). For MVPA, significant subgroup differences were observed by algorithm type (χ² = 8.22, df = 1, *p* = 0.004). Overall, when including all outcomes, subgroup differences were only statistically significant by comparison groups (χ² = 13.67, df = 4, *p* = 0.008). We further compared the effects of different chatbot features, as shown in Figure S3. Audio-based chatbots, chatbots using action planning, chatbots using visual cues, and humorous chatbot outputs showed larger effects than text-based chatbots, chatbots using coping planning, chatbots using relational cues, and non-humorous chatbot outputs, respectively. However, these differences were not statistically significant (*p* > 0.05).

Sensitive analysis including only 19 RCTs showed similar effects on overall effects, daily steps, MVPA, walking minutes, exercise time, and PA goal achievement compared with the overall post-intervention analysis (Figure S4). The only large difference was observed for total PA, which was significant in the overall analysis (g = 0.44, 95% CI [0.06, 0.82]) but not in the RCT-only analysis (g = 0.29, 95% CI [-0.57, 1.16]).

#### Follow-up effects

Figure 3 presents the follow-up effects of chatbot interventions on PA, including both RCTs and pre–post studies. Only a small number of studies (n = 4) provided evidence on post-intervention follow-up effects. Overall, chatbot interventions showed a small and non-statistically significant follow-up effect on PA (g = 0.10, 95% CI [-0.02, 0.21]). The sensitivity analysis including only RCTs for follow-up effects is similar (g = 0.11, 95% CI [-0.01, 0.22]) (see Figure S5).

**Figure 3.**
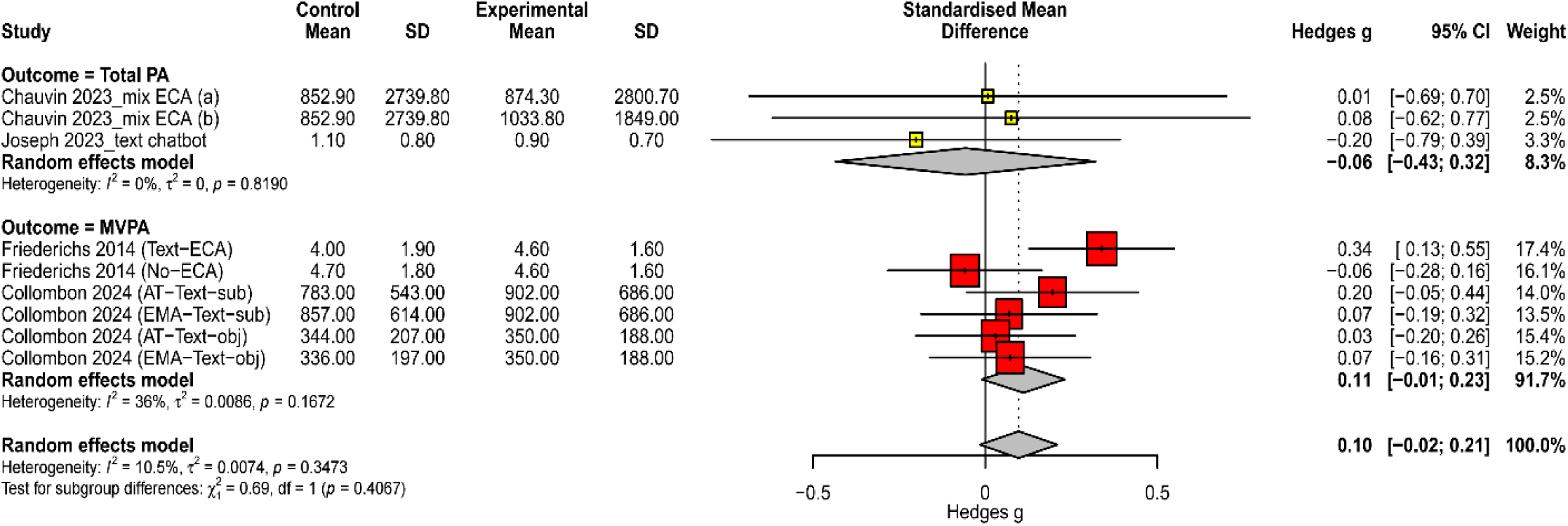
Follow-up forest plot by outcome ***Note*.** a: directed motivational interviewing; b: empathetic motivational interviewing; Text: text-based chatbot; AT: activity tracker group; EMA: ecological momentary assessment group; sub: subjective assessment; obj: objective assessment; No: no intervention received.

#### Publication bias

Funnel plots were examined separately for post-intervention (see Figure S6A) and follow-up studies (see Figure S6B). Although the post-intervention plot suggested some asymmetry, Egger’s test was not significant (bias = 1.27, SE = 1.11, t = 1.15, *p* = 0.260). This suggested no strong evidence of publication bias. Trim-and-fill identified three potentially missing studies, but the adjusted pooled effect remained significant (g = 0.31, 95% CI [0.14, 0.48]), indicating that the findings were relatively robust to potential publication bias. For the follow-up studies, Egger’s regression test was not conducted because the number of studies was too small to assess small-study effects. The trim-and-fill method identified three potentially missing studies. After adjustment, the pooled effect estimate changed but remained small and statistically significant (g = 0.11, 95% CI [0.03, 0.20]). These findings suggest that the follow-up effect may be influenced by potential publication bias and should be interpreted with caution.

### Meta-regression results

We included studies reporting daily step outcomes (n = 12) in the primary meta-regression analysis (see Table 1), because this is the only outcome that has more than 10 studies [47]. However, the multiple intervention moderator was not examined because only one study had no additional intervention. Categories represented by a single study, including no intervention, human advisor, and community setting, were also excluded. As a sensitivity analysis, we examined all post-intervention PA outcomes, including steps, MVPA, total PA, walking minutes, exercise time, and PA goal achievement (n = 24; see Table S7). The human advisor group was excluded because it included only one study. One study lacked setting data, and two lacked clinical-status data.

For step outcomes, intervention duration (QM = 6.09, *p* = 0.048) and participants’ clinic status (QM = 5.69, *p* = 0.017) were significant moderators. Interventions lasting 6–12 weeks showed a significantly stronger effect than those lasting less than 6 weeks (*β* = 0.62, *p* = 0.015), whereas interventions lasting more than 12 weeks did not show a significant advantage (*β* = 0.01, *p* = 0.962). Chatbot interventions showed significantly stronger effects among clinical populations than among non-clinical populations (*β* = 0.41, *p* = 0.017). Although other moderators were not statistically significant, several patterns were observed. The chatbot showed a smaller effect when compared with the chatbot base condition than when compared with usual care. However, this difference was not statistically significant (*β* = -0.35, *p* = 0.126). Compared with home-based intervention, free-living interventions showed relatively smaller effects (*β* = -0.09, *p* = 0.533). Chatbot interventions with embodied visual representations showed slightly smaller effects than without embodied conversational agents (*β* = -0.03, *p* = 0.849). However, because embodied conversational agents were more likely to be non-AI based and to use mixed output modalities, we conducted a multivariable meta-regression adjusting for output type and algorithm type. In the adjusted model, chatbot interventions without embodied visual representations continued to show a slightly larger effect than embodied conversational agents (*β* = 0.12, *p* = 0.650). Furthermore, audio-based chatbots showed slightly larger effects than text-based chatbots (*β* = 0.26, *p* = 0.266). Traditional AI-based chatbots showed slightly larger effects than non-AI chatbots (*β* = 0.21, *p* = 0.130), whereas generative AI-based chatbots were not represented in analyses of steps.

**Table 1.**
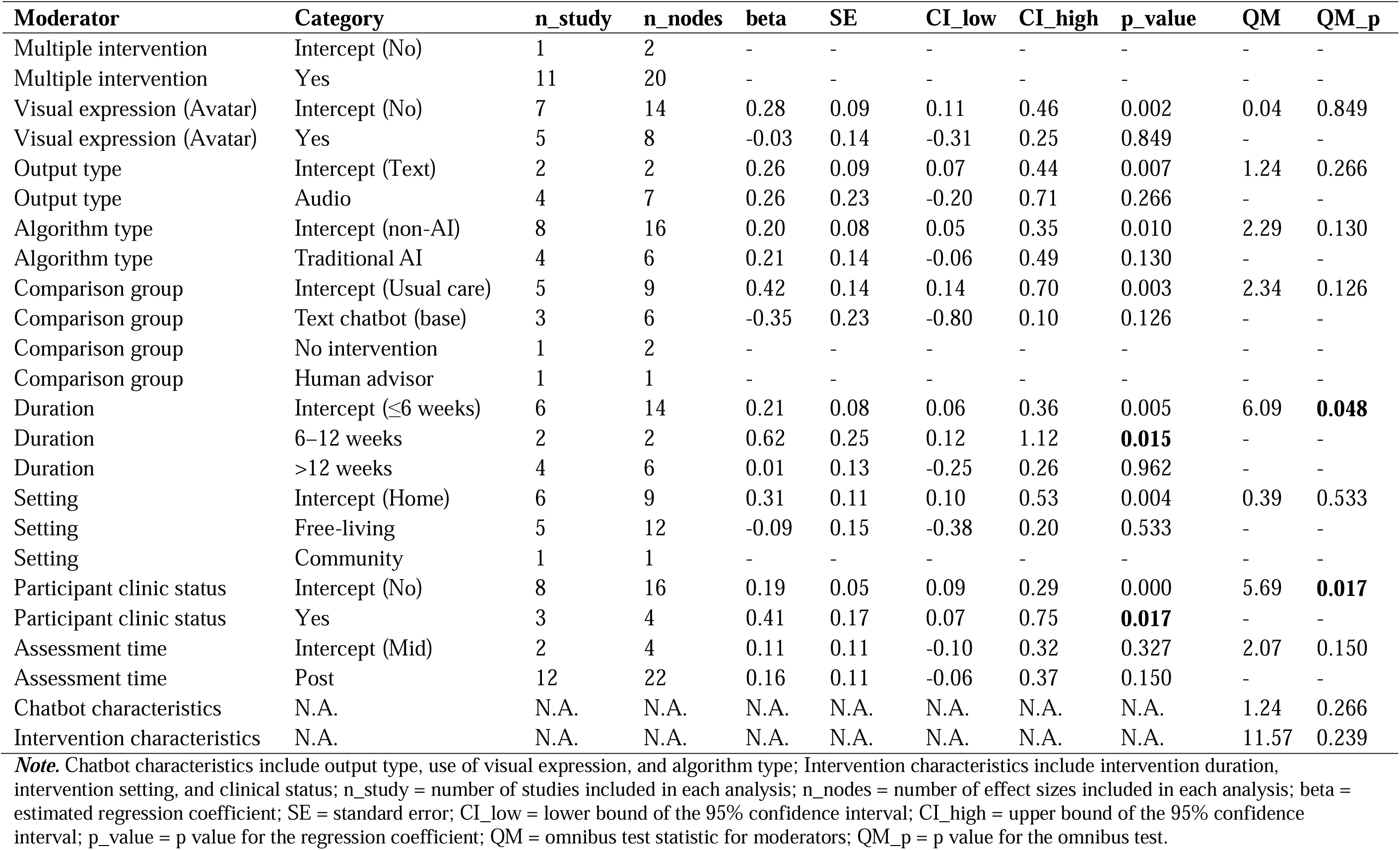
Meta-regression analysis (daily steps outcome)

| **Moderator** | **Category** | **n_study** | **n_nodes** | **beta** | **SE** | **CI_low** | **CI_high** | **p_value** | **QM** | **QM_p** |
|---|---|---|---|---|---|---|---|---|---|---|
| Multiple intervention | Intercept (No) | 1 | 2 | - | - | - | - | - | - | - |
| Multiple intervention | Yes | 11 | 20 | - | - | - | - | - | - | - |
| Visual expression (Avatar) | Intercept (No) | 7 | 14 | 0.28 | 0.09 | 0.11 | 0.46 | 0.002 | 0.04 | 0.849 |
| Visual expression (Avatar) | Yes | 5 | 8 | -0.03 | 0.14 | -0.31 | 0.25 | 0.849 | - | - |
| Output type | Intercept (Text) | 2 | 2 | 0.26 | 0.09 | 0.07 | 0.44 | 0.007 | 1.24 | 0.266 |
| Output type | Audio | 4 | 7 | 0.26 | 0.23 | -0.20 | 0.71 | 0.266 | - | - |
| Algorithm type | Intercept (non-AI) | 8 | 16 | 0.20 | 0.08 | 0.05 | 0.35 | 0.010 | 2.29 | 0.130 |
| Algorithm type | Traditional AI | 4 | 6 | 0.21 | 0.14 | -0.06 | 0.49 | 0.130 | - | - |
| Comparison group | Intercept (Usual care) | 5 | 9 | 0.42 | 0.14 | 0.14 | 0.70 | 0.003 | 2.34 | 0.126 |
| Comparison group | Text chatbot (base) | 3 | 6 | -0.35 | 0.23 | -0.80 | 0.10 | 0.126 | - | - |
| Comparison group | No intervention | 1 | 2 | - | - | - | - | - | - | - |
| Comparison group | Human advisor | 1 | 1 | - | - | - | - | - | - | - |
| Duration | Intercept (≤6 weeks) | 6 | 14 | 0.21 | 0.08 | 0.06 | 0.36 | 0.005 | 6.09 | **0.048** |
| Duration | 6–12 weeks | 2 | 2 | 0.62 | 0.25 | 0.12 | 1.12 | **0.015** | - | - |
| Duration | >12 weeks | 4 | 6 | 0.01 | 0.13 | -0.25 | 0.26 | 0.962 | - | - |
| Setting | Intercept (Home) | 6 | 9 | 0.31 | 0.11 | 0.10 | 0.53 | 0.004 | 0.39 | 0.533 |
| Setting | Free-living | 5 | 12 | -0.09 | 0.15 | -0.38 | 0.20 | 0.533 | - | - |
| Setting | Community | 1 | 1 | - | - | - | - | - | - | - |
| Participant clinic status | Intercept (No) | 8 | 16 | 0.19 | 0.05 | 0.09 | 0.29 | 0.000 | 5.69 | **0.017** |
| Participant clinic status | Yes | 3 | 4 | 0.41 | 0.17 | 0.07 | 0.75 | **0.017** | - | - |
| Assessment time | Intercept (Mid) | 2 | 4 | 0.11 | 0.11 | -0.10 | 0.32 | 0.327 | 2.07 | 0.150 |
| Assessment time | Post | 12 | 22 | 0.16 | 0.11 | -0.06 | 0.37 | 0.150 | - | - |
| Chatbot characteristics | N.A. | N.A. | N.A. | N.A. | N.A. | N.A. | N.A. | N.A. | 1.24 | 0.266 |
| Intervention characteristics | N.A. | N.A. | N.A. | N.A. | N.A. | N.A. | N.A. | N.A. | 11.57 | 0.239 |

***Note.*** Chatbot characteristics include output type, use of visual expression, and algorithm type; Intervention characteristics include intervention duration, intervention setting, and clinical status; n\_study = number of studies included in each analysis; n\_nodes = number of effect sizes included in each analysis; beta = estimated regression coefficient; SE = standard error; CI\_low = lower bound of the 95% confidence interval; CI\_high = upper bound of the 95% confidence interval; p\_value = p value for the regression coefficient; QM = omnibus test statistic for moderators; QM\_p = p value for the omnibus test.

In the sensitivity analysis including all PA outcomes, intervention duration and participants’ clinical status were no longer statistically significant moderators. Similar to the findings for step outcomes, all other moderators were not statistically significant. In contrast to step-only analysis, interventions lasting more than 12 weeks showed larger effects (*β* = 0.16, *p* = 0.411), as did interventions lasting 6–12 weeks (*β* = 0.13, *p* = 0.480), compared with interventions lasting less than 6 weeks. The broader analysis showed that chatbots with visual representations had larger, though non-significant, effects than non-visual chatbots (*β* = 0.09, *p* = 0.578). Complementing the step-only analysis, multi-component interventions showed slightly smaller, non-significant effects than chatbot-only interventions (*β* = -0.17, *p* = 0.500). Chatbots showed smaller effects when compared with no intervention than when compared with usual care (*β* = -0.38, *p* = 0.291), digital non-chatbot interventions (*β* = -0.88, *p* = 0.029), and text chatbot baseline groups (*β* = -0.73, *p* = 0.076). Studies conducted in home settings (*β* = -0.50, *p* = 0.052) and free-living settings (*β* = -0.43, *p* = 0.072) showed smaller effects than those conducted in community settings. Regarding output type, mixed-output chatbots showed larger, though non-significant, effects than text-only chatbots (*β* = 0.15, *p* = 0.416). Traditional AI-based chatbots showed relatively larger effects than non-AI chatbots (*β* = 0.18, *p* = 0.266), whereas generative AI-based chatbots showed smaller, non-significant effects compared with non-AI chatbots (*β* = 0.11, *p* = 0.672). We also examined whether intervention effects differed by assessment time point in the broader analyses. Compared with mid-intervention assessments, post-intervention assessments showed a slight increase in PA (*β* = 0.13, *p* = 0.382), whereas follow-up assessments showed a slight decrease (*β* = -0.04, *p* = 0.846).

## Discussion

Overall, AI chatbot interventions demonstrated statistically significant small-to-moderate efficacy in promoting physical activity (PA) across different outcomes at post-intervention. This finding is consistent with previous meta-analyses of chatbot interventions for PA [11,12]. The observed effects are also comparable to those reported in other digital health interventions for PA [48,49] and a benchmark effect size (Cohen’s d = 0.21) for PA interventions reported by Wright et al. (2021) [50]. However, this finding should be interpreted cautiously because most studies included in the meta-analysis were rated as having some concerns regarding risk of bias. When different outcomes were considered, significant changes were observed only for walking minutes, total PA, and daily steps, with effect sizes decreasing in that order. Chatbots showed only a small, non-significant effect on MVPA. These findings differ from Singh et al. (2023) [11], who reported stronger effects for MVPA than for total PA and steps, but is consistent with Bi et al. (2024) [51], who found stronger effects on steps than MVPA. One possible explanation is that walking minutes, total PA, and steps capture a broader range of movement, including light-intensity PA as well as MVPA. In contrast, increasing MVPA generally requires individuals to engage in activity at a higher intensity (>100 steps/minutes) [52], such as brisk walking or structured exercise, which may be more difficult to achieve and maintain. Therefore, chatbot interventions may be more effective in promoting overall movement or step accumulation than in increasing MVPA. When comparing walking, total PA, and step counts, the walking minutes and total PA outcomes included in this meta-analysis were primarily assessed using self-reported approach. In contrast, step counts were generally assessed objectively. It is well known that participants may overreport their PA levels when conducting self-report surveys. This issue may be more pronounced in digital intervention studies, where participants often cannot be blinded to the intervention conditions. Therefore, walking minutes and total PA showed greater improvement than step counts after the chatbot intervention. However, researchers should be cautious that the results for walking minutes were only from two non-RCTs [53,54]. Moreover, RCTs-only sensitivity analysis showed a non-significant effect for total PA, with an effect size relatively like that observed for steps. This means more RCTs are needed to explore the efficacy on walking minutes and total PA.

A small, non-significant effect size was observed at follow-up assessments. This provides new synthesized evidence compared with previous meta-analyses [11,12]. The small follow-up effect size is consistent with findings from other digital health interventions targeting PA, which reported a pooled effect of SMD = 0.17 (95% CI: 0.04–0.31) [48]. However, the follow-up effect should be interpreted with caution because it was based on only four studies. In the meta-regression analysis including all PA outcomes, a similar pattern was observed, with follow-up effects being slightly smaller than post-intervention effects when compared with mid-intervention effects. However, assessment time did not significantly moderate the effects of chatbot interventions on PA outcomes. One potential concern regarding the meta-regression of assessment time is the uneven distribution of studies across assessment points, with most studies provided post-intervention evidence, whereas relatively few studies examined mid-intervention and follow-up effects. Therefore, we were unable to draw a firm conclusion about whether intervention effects are maintained over the long term after chatbot interventions. Future studies are encouraged to provide more evidence on follow-up effects. In addition, because of the limited number of studies evaluating follow-up outcomes, we were also unable to examine whether factors such as intervention duration, chatbot characteristics, and user engagement moderate the long-term maintenance of PA after chatbot interventions. For example, future studies could examine whether longer intervention duration helps maintain PA engagement after the intervention ends.

The exploratory subgroup analyses suggested that chatbot effects may differ by comparison group type, such as usual care, human advisor, or digital non-chatbot controls, particularly for total PA, exercise time, and all PA outcomes. However, in the meta-regression analyses including daily steps or all PA outcomes, comparison group type was not a statistically significant moderator. This discrepancy may reflect the difference between descriptive subgroup comparisons and formal moderator testing, especially after accounting for dependency among effect sizes. Therefore, although comparison group type may contribute to heterogeneity for some specific outcomes, it should not be interpreted as a robust or consistent moderator of chatbot intervention effects. In particular, chatbot interventions were associated with greater increases in daily steps compared with usual care in both the subgroup and meta-regression analyses. One possible explanation is that participants in the chatbot intervention groups often continued to receive usual care, with the chatbot added as an additional PA support component. Therefore, the observed effects may reflect the added benefit of chatbot-based support beyond traditional usual care. Finally, only one study compared chatbots with human advisors,[54] limiting conclusions about their relative effectiveness. Further comparative research is needed.

When intervention duration, participants’ clinical status, and intervention setting were examined together as intervention characteristics, the combined model did not show a statistically significant moderating effect. Similarly, subgroup analyses suggested that the effects of chatbot interventions on daily steps did not differ by intervention duration, which is consistent with another meta-analysis comparing interventions lasting ≤6 weeks versus >6 weeks [11]. However, in the meta-regression analysis of daily step outcomes, intervention duration was a significant moderator. Specifically, interventions lasting 6–12 weeks had significantly greater effects on daily steps than those lasting ≤6 weeks, whereas interventions lasting >12 weeks showed smaller, non-significant effects compared with those lasting ≤6 weeks. The reduced effects observed in longer-duration interventions are consistent with Wang et al. (2025) [12], who found that chatbot interventions produced a modest but statistically significant increase in PA over shorter durations (≤8 weeks), but not over longer durations (>8 weeks). Although we did not have sufficient data to examine whether the association between engagement and PA outcomes changed over time, some studies found that higher engagement (i.e., usage measures) was associated with greater PA participation [41,55]. Therefore, the lower effects observed after longer intervention durations may be partly explained by reduced adherence or engagement over time, which could weaken post-intervention effects. However, these findings should be interpreted cautiously because the number of studies was unevenly distributed across duration categories, with 50% of studies lasting less than 6 weeks. In addition, when all PA outcomes were included in the sensitivity meta-analysis, intervention duration was no longer a significant moderator. This may be because including multiple PA outcomes increased heterogeneity in measurement tools and units, thereby increasing uncertainty around the moderator effect. Future studies should provide stronger evidence on the moderating role of intervention duration across different outcomes, such as MVPA and total PA, and further examine how engagement patterns change over time and relate to post-intervention efficacy. Building on previous reviews [11,12], we further found a significant moderating effects of clinical status on daily steps, but not on across all outcome meta-regression. Clinical participants showed a significantly greater increase in daily steps compared with non-clinical participants. This might be because participants in clinical samples may have greater perceived needs, clearer health goals, and stronger intuitive motivation, which could enhance responsiveness to chatbot-based support. However, the number of studies was unevenly distributed between clinical (n = 3) and non-clinical (n = 8) populations. Additional studies are needed to confirm these findings. Furthermore, intervention setting was not a statistically significant moderator. However, the step-specific meta-regression suggested slightly smaller effects in free-living settings compared with home-based settings. In the meta-regression including all PA outcomes, home and free-living settings showed non-significantly smaller effects than more structured community settings. These findings suggested that chatbot interventions may have stronger effects in more controlled or structured settings. Future research should further explore strategies to enhance intervention effects in free-living settings.

Regarding chatbot characteristics, the multivariate models including output type, use of visual expression, and algorithm type did not show statistically significant moderating effects. In addition, neither chatbot output type nor algorithm type significantly moderated intervention effects in either the step-specific or the all PA outcomes meta-regression, which is consistent with previous meta-analytic findings [11]. Some interesting patterns were observed. Only the meta-regression including all PA outcomes had sufficient data to examine differences among generative AI, traditional AI, and non-AI chatbots. The results showed that both traditional AI and generative AI chatbots had non-significantly larger effects than non-AI chatbots. Traditional AI chatbots also showed larger effects than generative AI chatbots relative to non-AI chatbots. However, these findings should be interpreted cautiously because only three studies used generative AI approaches, which may have limited statistical power. More evidence is needed to compare these approaches in chatbot interventions. Moreover, we did not find a statistically significant effect of visual representation, such as embodied conversational agents, compared with chatbots without visual representation. One possible explanation is that most chatbots with visual representation used non-AI algorithms, which may have limited the potential benefits of visual embodiment. Future studies should explore whether combining visual representation with AI algorithms can enhance the effectiveness of chatbots in promoting PA. We also examined different chatbot features and found several non-significant differences. For example, audio-based chatbots [35], action planning [56], humorous outputs[57], and visual cues [58] showed slightly larger effects than their comparison features, including text-based chatbots, coping planning, non-humorous outputs, and relational cues. These findings suggested that modifying a single chatbot feature may not be sufficient to produce substantial benefits. Future studies should test combinations of features to better understand how chatbot design can be optimized for promoting PA.

### Contribution, limitations, and implications

This review used rigorous methods, including searches of 11 databases, multistage screening, risk-of-bias assessment, comprehensive data extraction, meta-analysis, and sensitivity analyses. It extended previous subgroup analyses [11,12] by using meta-regression to formally test multiple participant-, intervention-, and chatbot-related moderators, although findings should be interpreted cautiously because of the limited number of studies. It also allows multiple moderators to be tested simultaneously. Second, this study provided preliminary evidence on the sustainability of chatbot effects by analyzing the follow-up effects. Third, analyses of intervention setting and clinical status suggested that chatbots may be useful across controlled and free-living settings and in clinical populations.

Several limitations should be acknowledged. First, only studies published in English were included, which may have introduced language bias. Second, the review deviated from the registered protocol by narrowing the outcomes to physical activity and expanding the planned moderators from five to 11. Finally, the certainty of the overall body of evidence was not formally assessed using a framework such as the Grading of Recommendations Assessment, Development and Evaluation (GRADE) approach. The study’s conclusions are also limited by the current evidence base. Most studies included in the meta-analysis were rated as having some concerns regarding risk of bias. Most studies included predominantly female adults from non-clinical populations and evaluated text-based chatbots within multicomponent interventions. Voluntary recruitment may also have underrepresented underserved individuals and those with lower motivation or literacy, limiting generalizability. Although randomization supports causal interpretation of overall intervention effects, subgroup and meta-regression analyses rely on nonrandomized study-level characteristics. These analyses may identify potential sources of heterogeneity but cannot establish causal moderation.

Evidence suggests that chatbots are promising tools for promoting PA in clinical and non-clinical populations. Future research should prioritize rigorous randomized controlled trials with larger samples (>100), longer interventions (>12 weeks), and follow-up assessments. More studies are also needed among adolescents, sex-diverse populations, and clinical groups. Researchers should also examine changes in behavioral and emotional engagement over time and their associations with post-intervention and follow-up intervention outcomes. Direct comparisons with human advisors and non-chatbot digital interventions are needed to establish the added value of chatbots. Such comparisons are necessary to ensure that claims regarding the added value of chatbots are supported by evidence rather than based primarily on theoretical assumptions. Future studies should also compare generative and traditional AI chatbots, evaluate models beyond GPT-based systems, and test combinations of design features, including avatar-based interfaces, empathy, dialogue flexibility, and humor.

## Conclusions

This meta-analysis found statistically significant small-to-moderate effects of chatbot interventions on PA. Several subgroup differences across PA outcomes helped explain heterogeneity. In the meta-regression analyses, intervention duration and participants’ clinical status were significant moderators of daily steps. Future large-scale trials with rigorous designs are needed, along with additional studies to provide sufficient power to determine how participant, intervention, and chatbot characteristics influence the effectiveness of chatbots for PA promotion.

## Data Availability

All data produced in the present study are available upon reasonable request to the authors.

## Notes

### Competing Interest Statement

The authors have declared no competing interest.

