## Supplementary Figure S1-S5 for "Chatbot as an intervention on physical activity: A systematic review and meta-analysis with meta-regression"

**Supplementary Materials**

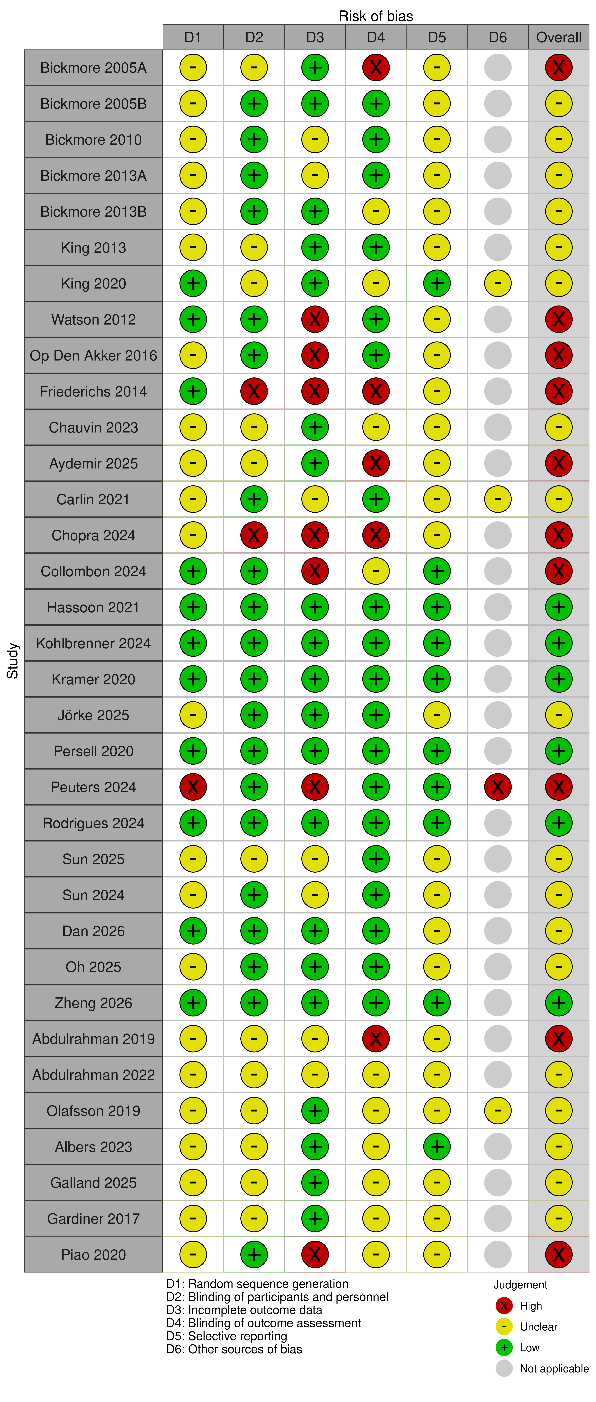

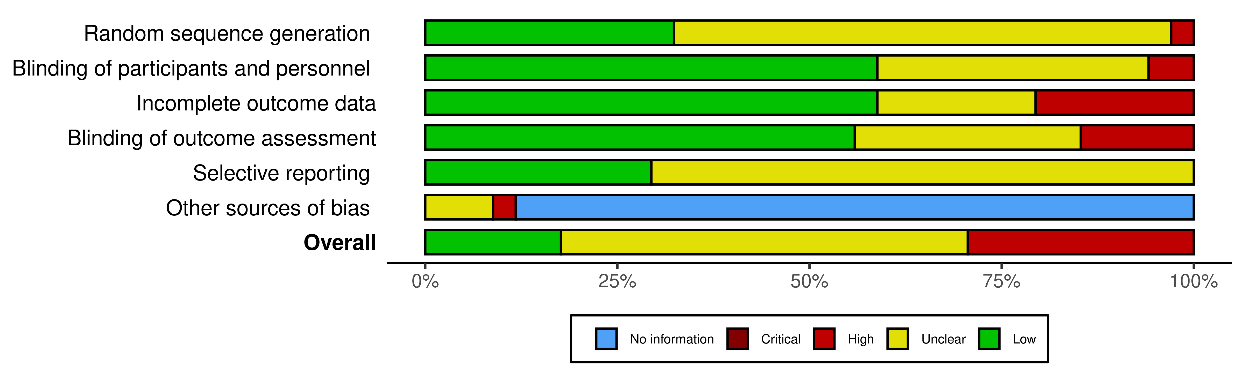

**A**

**B**

**Figure S1.** Risk of bias in randomized controlled trials (RCTs)

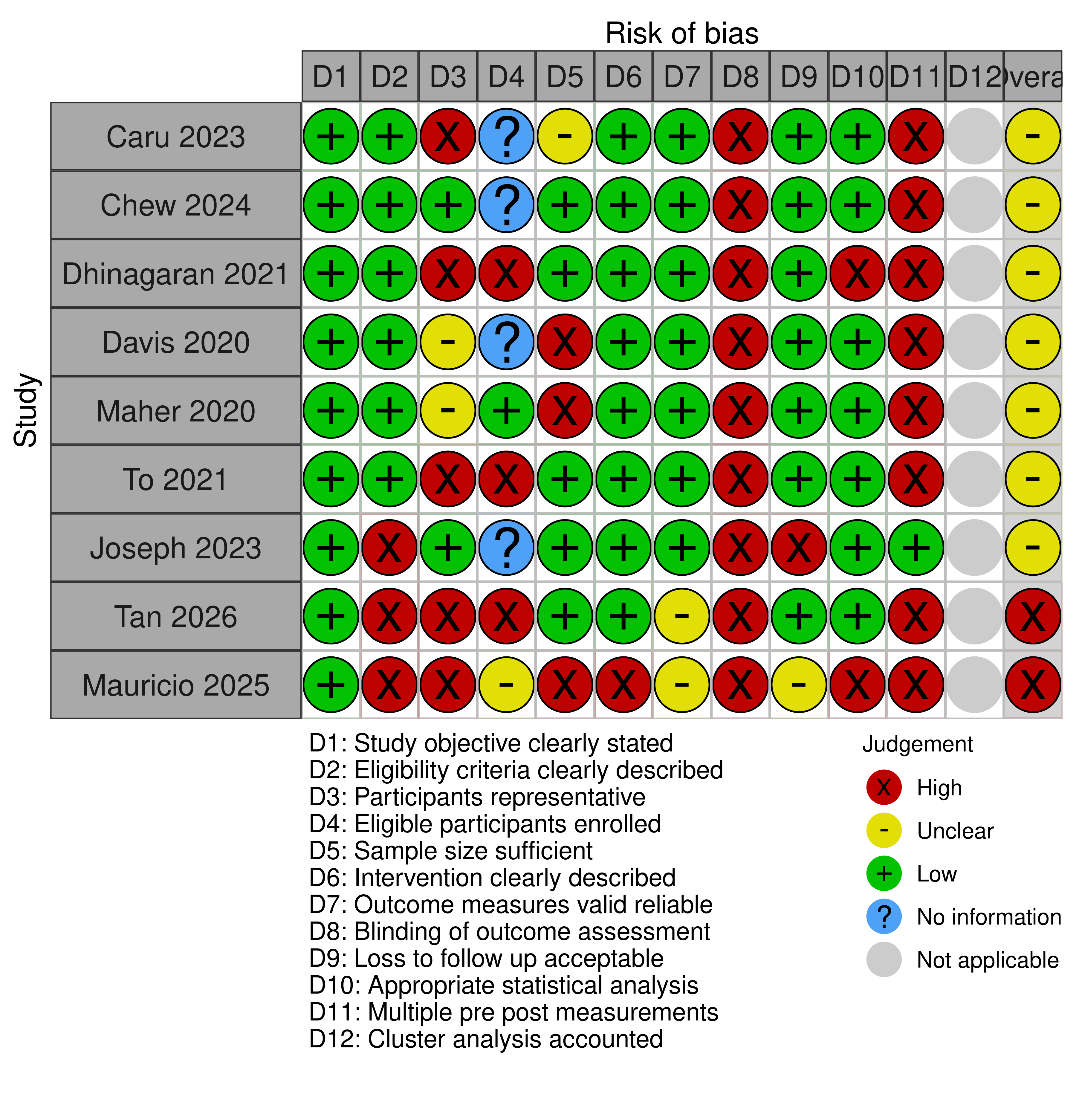

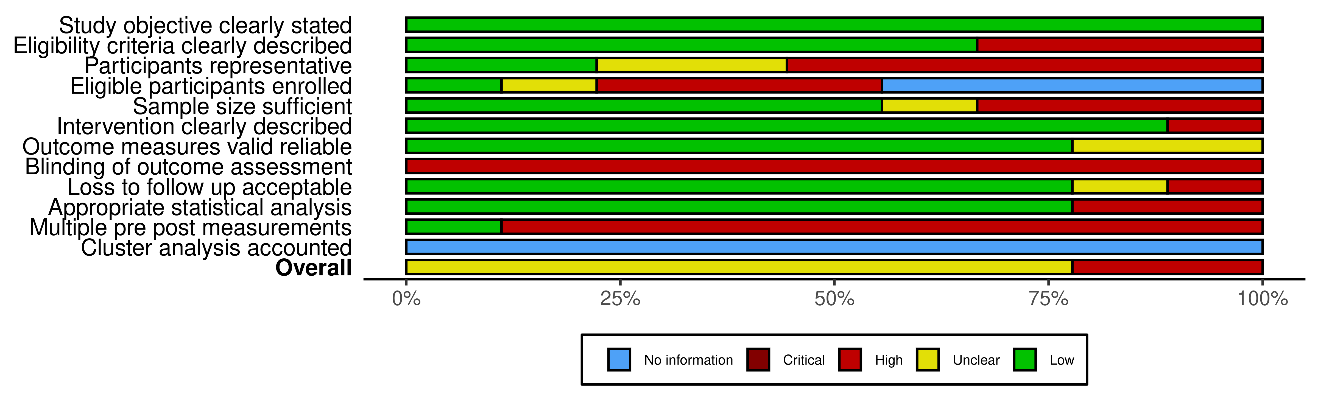

**B**

**A**

**Figure S2.** Risk of bias in pre-post studies

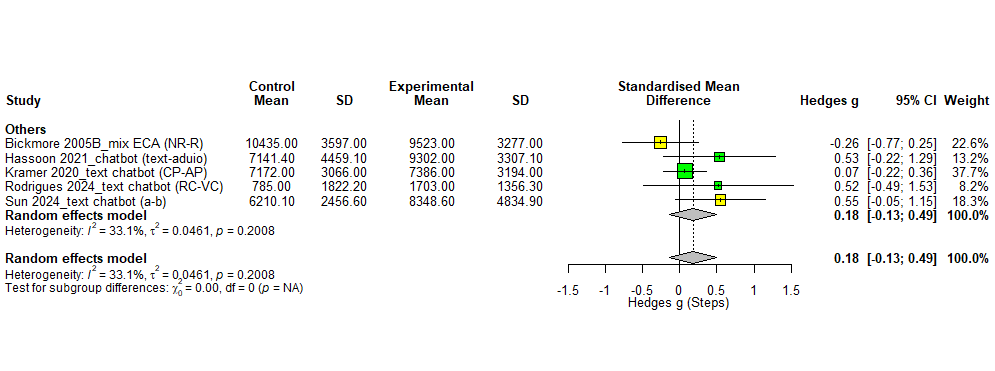

**Figure S3.** Comparison of chatbot features (RCTs and pre-post studies)

***Note*.** NR: non-relational; R: relational; text: text-based chatbot; audio: audio-based chatbot; CP: coping planning; AP: action planning; RC: relational cues; VC: visual cues; a: non-humorous; b: humorous.

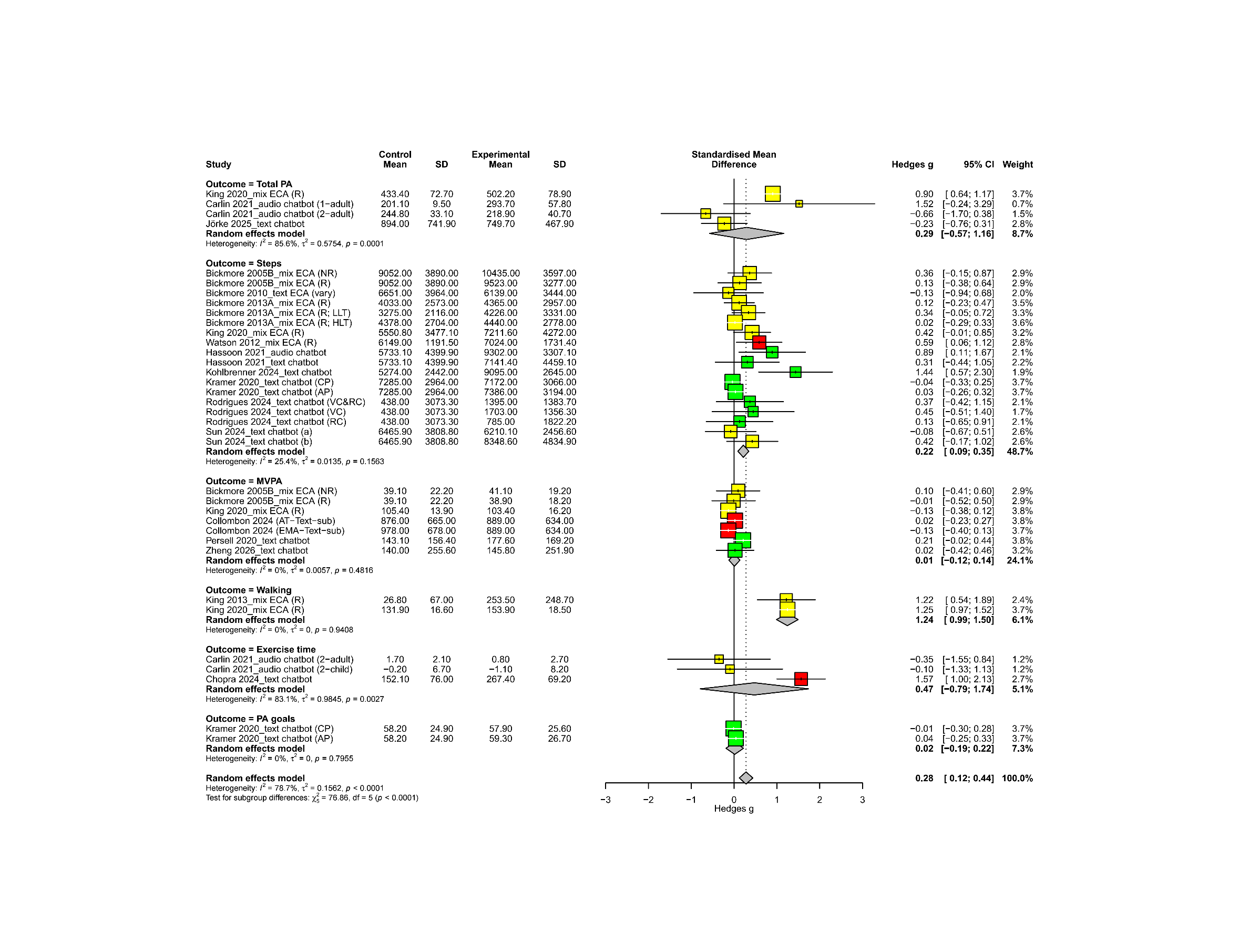

**Figure S4.** Post-intervention forest plot by outcome (RCTs)

***Note*.** Mix: chatbots that used both text- and audio-based communication; 1-adult: phase 1 data for adults; 2-adult: phase 2 data for adult; 2-child: phase 2 data for children; NR: non-relational; R: relational; LLT: low-literacy population; HLT: high-literacy population; Vary: dialogue structures changed in each conversation; CP: coping planning; AP: action planning; VC: visual cues; RC: relational cues; HC: human cues; a: non-humorous; b: humorous; AT: activity tracker; EMA: ecological momentary assessment; Text: text-based chatbot; sub: subjective assessment.

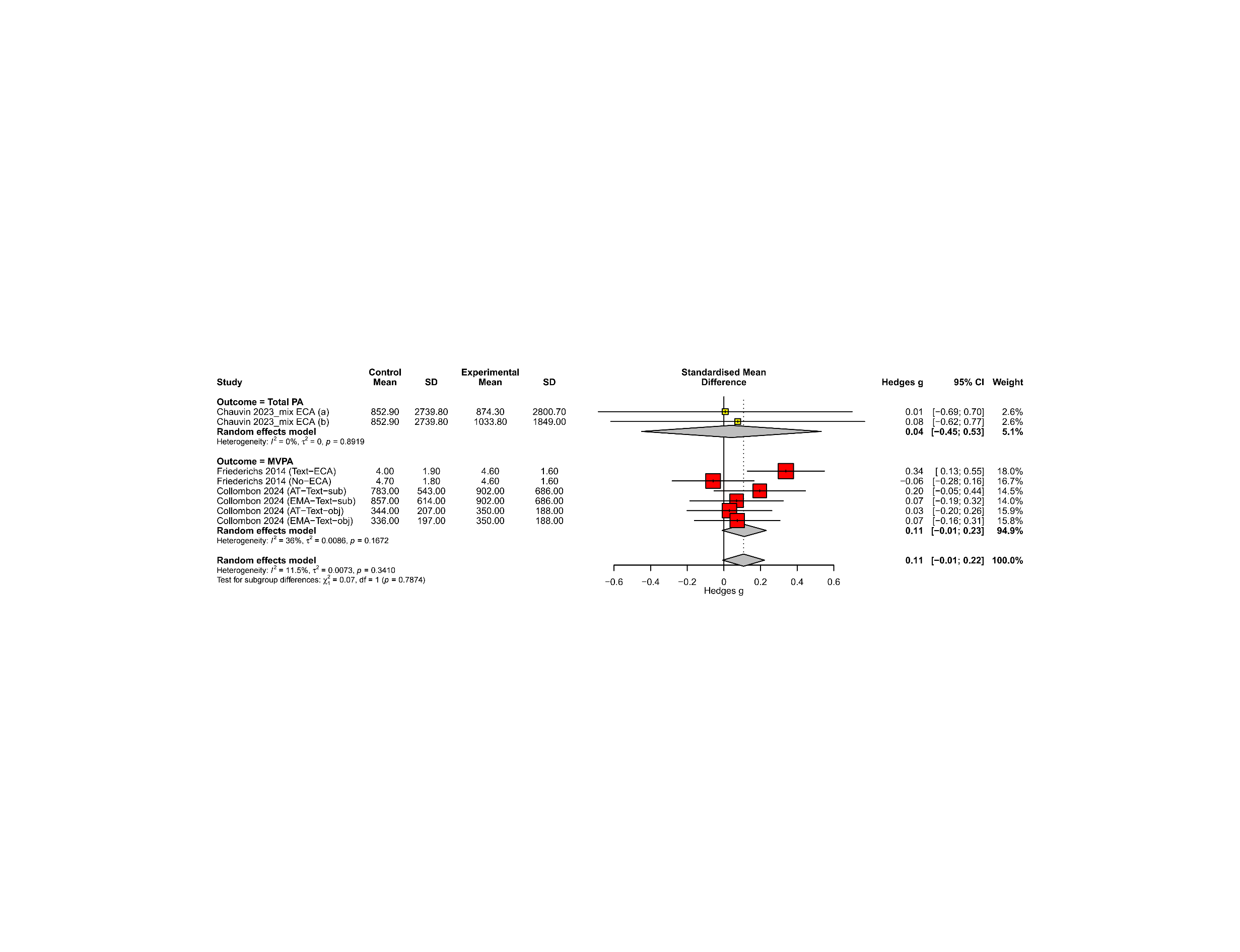

**Figure S5.** Follow-up forest plot by outcome (RCTs)

***Note.*** a: directed motivational interviewing; b: empathetic motivational interviewing
