## Supplementary Table S1-S6 for "Chatbot as an intervention on physical activity: A systematic review and meta-analysis with meta-regression"

**Table S1.** Search Terms Used in Each Database

**Web of Science (.ris)**

| **Search date** | 1/27/2026 |
| --- | --- |
| **Search terms** | (TS=(lifestyle* OR "health behavior*" OR exercise* OR "physical activ*") AND TS=(Chatbot* OR chat-bot* OR "chat bot*" OR "social bot*" OR eCoach* OR infobot* OR smartbot* OR "conversational bot*" OR "artificial intelligence chatbot*" OR "AI agent*" OR "conversational agent*" OR "dialogue agent*"OR "dialog agent*" OR "artificial agent" OR "interactive agent*" OR "virtual agent*" OR "automated agent*" OR "relational agent*" OR "dialogue agent*" OR "dialog agent*" OR "AI assist*" OR "virtual assist*" OR "conversational assistant*" OR "digital assist*" OR "intelligent assist*" OR "virtual assist*" OR "smart assist*" OR "virtual health assist*" OR "AI advisor*" OR "virtual advisor*" OR "animated advisor*" OR "smart advisor*" OR "AI coach*" OR "virtual coach*" OR "smart coach*" OR "animated coach*" OR "artificial conversation entit*" OR "Assistance technolog*" OR "conversational AI" OR "conversational interface*" OR "conversational system*" OR "dialogue system*" OR "natural language interface*" OR "automated conversation" OR "virtual conversation" OR ChatGPT OR Llama OR DeepSeek)) |
| **Refine** | Languages = English |
|  | Document Types = Article or Early Access, or Proceeding Paper |
| **Number** | **Original search scope: Physical activity, sleep, and sedentary**   - Records identified in the original search: N = 1,391 (6/18/2025) - Additional records identified: N = 13 (7/1/2025) - Additional records identified: N = 2 (10/25/2025)   **After protocol revision: Physical activity only**   - Updated total records identified: N = 1,665 (1/27/2026) - Newly added records after duplicate removal: N = 539 (1/27/2026) |
| **Note** | Search by Topic (LS)  **Early access:** An article that has been electronically published by a journal before it has been assigned to a specific volume and issue.  **Proceedings paper:** Full papers in a wide range of disciplines that were or will be presented at a symposium or meeting. The papers to be included must have been presented in full at a conference, meeting, symposium, or similar gathering. Generally published in a book of conference proceedings. |

**PubMed (.nbib)**

| **Search date** | 1/27/2026 |
| --- | --- |
| **Search terms** | (((lifestyle*[tiab] OR health behavior*[tiab] OR exercise*[tiab] OR physical activ*[tiab]) AND (Chatbot*[tiab] OR chat-bot*[tiab] OR chat bot*[tiab] OR social bot*[tiab] OR eCoach[tiab]* OR infobot*[tiab] OR smartbot*[tiab] OR conversational bot*[tiab] OR artificial intelligence chatbot*[tiab] OR AI agent*[tiab] OR conversational agent*[tiab] OR dialogue agent*[tiab] OR dialog agent*[tiab] OR artificial agent[tiab] OR interactive agent*[tiab] OR virtual agent*[tiab] OR automated agent*[tiab] OR relational agent*[tiab] OR dialogue agent*[tiab] OR dialog agent*[tiab] OR AI assist*[tiab] OR virtual assist*[tiab] OR conversational assistant*[tiab] OR digital assist*[tiab] OR intelligent assist*[tiab] OR virtual assist*[tiab] OR smart assist*[tiab] OR virtual health assist*[tiab] OR AI advisor*[tiab] OR virtual advisor*[tiab] OR animated advisor*[tiab] OR smart advisor*[tiab] OR AI coach*[tiab] OR virtual coach*[tiab] OR smart coach*[tiab] OR animated coach*[tiab] OR artificial conversation entit*[tiab] OR Assistance technolog*[tiab] OR conversational AI*[tiab] OR conversational interface*[tiab] OR conversational system*[tiab] OR dialogue system*[tiab] OR natural language interface*[tiab] OR automated conversation*[tiab] OR virtual conversation*[tiab] OR ChatGPT*[tiab] OR Llama*[tiab] OR DeepSeek*[tiab])) AND (eng[la])) NOT (Systematic review[pt] OR meta-analysis[pt] OR review[pt]) |
| **Numbers** | **Original search scope: Physical activity, sleep, and sedentary**   - Records identified in the original search: N = 622 (6/18/2025) - Additional records identified: N = 56 (7/1/2025)   **After protocol revision: Physical activity only**   - Updated total records identified: N = 657 (1/27/2026) - Newly added records after duplicate removal: N = 28 (1/27/2026) |
| **Note** | Search by title and abstract [tiab]  Language type [la]  Publication type [pt] |

***CINAHL (Cumulative Index to Nursing and Allied Health Literature) (.ris)***

| **Search date** | **1/27/2026** |
| --- | --- |
| **Search terms** | XB ( (lifestyle* OR "health behavior*" OR exercise* OR "physical activ*") AND XB (Chatbot* OR chat-bot* OR "chat bot*" OR "social bot*" OR eCoach* OR infobot* OR smartbot* OR "conversational bot*" OR "artificial intelligence chatbot*" OR "AI agent*" OR "conversational agent*" OR "dialogue agent*"OR "dialog agent*" OR "artificial agent" OR "interactive agent*" OR "virtual agent*" OR "automated agent*" OR "relational agent*" OR "dialogue agent*" OR "dialog agent*" OR "AI assist*" OR "virtual assist*" OR "conversational assistant*" OR "digital assist*" OR "intelligent assist*" OR "virtual assist*" OR "smart assist*" OR "virtual health assist*" OR "AI advisor*" OR "virtual advisor*" OR "animated advisor*" OR "smart advisor*" OR "AI coach*" OR "virtual coach*" OR "smart coach*" OR "animated coach*" OR "artificial conversation entit*" OR "Assistance technolog*" OR "conversational AI" OR "conversational interface*" OR "conversational system*" OR "dialogue system*" OR "natural language interface*" OR "automated conversation" OR "virtual conversation" OR ChatGPT OR Llama OR DeepSeek ) AND LA English NOT PT ( Systematic review OR meta-analysis OR review ) |
| **Numbers** | **Original search scope: Physical activity, sleep, and sedentary**   - Records identified in the original search: N = 156 (6/18/2025)   **After protocol revision: Physical activity only**   - Updated total records identified: N = 161 (1/27/2026) - Newly added records after duplicate removal: N = 7 (1/27/2026) |
| **Note** | Search by title and abstract (XB) |

***Medline (.ris)***

| **Search date** | 1/27/2026 |
| --- | --- |
| **Search term**s | (TS=(lifestyle* OR "health behavior*" OR exercise* OR "physical activ*") AND TS=(Chatbot* OR chat-bot* OR "chat bot*" OR "social bot*" OR eCoach* OR infobot* OR smartbot* OR "conversational bot*" OR "artificial intelligence chatbot*" OR "AI agent*" OR "conversational agent*" OR "dialogue agent*"OR "dialog agent*" OR "artificial agent" OR "interactive agent*" OR "virtual agent*" OR "automated agent*" OR "relational agent*" OR "dialogue agent*" OR "dialog agent*" OR "AI assist*" OR "virtual assist*" OR "conversational assistant*" OR "digital assist*" OR "intelligent assist*" OR "virtual assist*" OR "smart assist*" OR "virtual health assist*" OR "AI advisor*" OR "virtual advisor*" OR "animated advisor*" OR "smart advisor*" OR "AI coach*" OR "virtual coach*" OR "smart coach*" OR "animated coach*" OR "artificial conversation entit*" OR "Assistance technolog*" OR "conversational AI" OR "conversational interface*" OR "conversational system*" OR "dialogue system*" OR "natural language interface*" OR "automated conversation" OR "virtual conversation" OR ChatGPT OR Llama OR DeepSeek)) |
| **Refine** | Languages = English |
|  | Document Types = Journal Article, Randomized controlled trail, comparative study, evaluation study |
| **Numbers** | **Original search scope: Physical activity, sleep, and sedentary**   - Records identified in the original search: N = 599 (6/18/2025)   **After protocol revision: Physical activity only**   - Updated total records identified: N = 667 (1/27/2026) - Newly added records after duplicate removal: N = 58 (1/27/2026) |
| **Note** | Search by title and abstract (XB) |

***EMBASE (Excerpta Medica dataBASE) (.ris)***

| **Search date** | 1/27/2026 |
| --- | --- |
| **Search terms** | (lifestyle*:ti,ab,kw OR 'health behavior*':ti,ab,kw OR exercise*:ti,ab,kw OR 'physical activ*':ti,ab,kw) AND (chatbot*:ti,ab,kw OR 'chat bot*':ti,ab,kw OR 'social bot*':ti,ab,kw OR eCoach*:ti,ab,kw OR infobot*:ti,ab,kw OR smartbot*:ti,ab,kw OR 'conversational bot*':ti,ab,kw OR 'artificial intelligence chatbot*':ti,ab,kw OR 'ai agent*':ti,ab,kw OR 'conversational agent*':ti,ab,kw OR 'artificial agent':ti,ab,kw OR 'interactive agent*':ti,ab,kw OR 'virtual agent*':ti,ab,kw OR 'automated agent*':ti,ab,kw OR 'relational agent*':ti,ab,kw OR 'dialogue agent*':ti,ab,kw OR 'dialog agent*':ti,ab,kw OR 'ai assist*':ti,ab,kw OR 'conversational assistant*':ti,ab,kw OR 'digital assist*':ti,ab,kw OR 'intelligent assist*':ti,ab,kw OR 'virtual assist*':ti,ab,kw OR 'smart assist*':ti,ab,kw OR 'virtual health assist*':ti,ab,kw OR 'ai advisor*':ti,ab,kw OR 'virtual advisor*':ti,ab,kw OR 'animated advisor*':ti,ab,kw OR 'smart advisor*':ti,ab,kw OR 'ai coach*':ti,ab,kw OR 'virtual coach*':ti,ab,kw OR 'smart coach*':ti,ab,kw OR 'animated coach*':ti,ab,kw OR 'artificial conversation entit*':ti,ab,kw OR 'assistance technolog*':ti,ab,kw OR 'conversational ai*':ti,ab,kw OR 'conversational interface*':ti,ab,kw OR 'conversational system*':ti,ab,kw OR 'dialogue system*':ti,ab,kw OR 'natural language interface*':ti,ab,kw OR 'automated conversation*':ti,ab,kw OR 'virtual conversation*':ti,ab,kw OR chatgpt*:ti,ab,kw OR llama*:ti,ab,kw OR deepseek*:ti,ab,kw) AND english:la NOT review:it |
| **Numbers** | **Original search scope: Physical activity, sleep, and sedentary**   - Records identified in the original search: N = 960 (6/18/2025)   **After protocol revision: Physical activity only**   - Updated total records identified: N = 1,056 (1/27/2026) - Newly added records after duplicate removal: N = 533 (1/7/2026) |
| **Note** | Search by title, abstract, and author keywords (ti,ab,kw)  Language (la)  Publication type (it) |

***Scopus (scientific, technical, medical and social sciences literature) (.ris)***

| **Search date** | 1/27/2026 |
| --- | --- |
| **Search syntax** | Advanced query  (  TITLE-ABS-KEY ( lifestyle* OR "health behavior*" OR exercise* OR "physical activ*" ) AND TITLE-ABS-KEY ( chatbot* OR chat-bot* OR "chat bot*" OR "social bot*" OR eCoach* OR infobot* OR smartbot* OR "conversational bot*" OR "artificial intelligence chatbot*" OR "AI agent*" OR "conversational agent*" OR "dialogue agent*" OR "dialog agent*" OR "artificial agent" OR "interactive agent*" OR "virtual agent*" OR "automated agent*" OR "relational agent*" OR "dialogue agent*" OR "dialog agent*" OR "AI assist*" OR "virtual assist*" OR "conversational assistant*" OR "digital assist*" OR "intelligent assist*" OR "virtual assist*" OR "smart assist*" OR "virtual health assist*" OR "AI advisor*" OR "virtual advisor*" OR "animated advisor*" OR "smart advisor*" OR "AI coach*" OR "virtual coach*" OR "smart coach*" OR "animated coach*" OR "artificial conversation entit*" OR "Assistance technolog*" OR "conversational AI" OR "conversational interface*" OR "conversational system*" OR "dialogue system*" OR "natural language interface*" OR "automated conversation" OR "virtual conversation" OR chatgpt OR llama OR deepseek ) AND LANGUAGE ( english ) ) AND ( LIMIT-TO ( DOCTYPE , "ar" ) OR LIMIT-TO ( DOCTYPE , "cp" ) ) |
| **Numbers** | **Original search scope: Physical activity, sleep, and sedentary**   - Records identified in the original search: N = 2,150 (6/18/2025)   **After protocol revision: Physical activity only**   - Updated total records identified: N = 2,210 (1/27/2025) - Newly added records after duplicate removal: N = 903 (1/27/2025) |
| **Note** | Search by Abstract title, Abstract, Keywords, Authors (TITLE-ABS-KEY-AUTH)  "ar": article  "cp": conference paper |

***APA PsycINFO (database produced by APA) (.ris)***

| **Search date** | 1/27/2026 |
| --- | --- |
| **Search terms** | XB (lifestyle* OR "health behavior*" OR exercise* OR "physical activ*" ) AND XB ( Chatbot* OR chat-bot* OR "chat bot*" OR "social bot*" OR eCoach* OR infobot* OR smartbot* OR "conversational bot*" OR "artificial intelligence chatbot*" OR "AI agent*" OR "conversational agent*" OR "dialogue agent*"OR "dialog agent*" OR "artificial agent" OR "interactive agent*" OR "virtual agent*" OR "automated agent*" OR "relational agent*" OR "dialogue agent*" OR "dialog agent*" OR "AI assist*" OR "virtual assist*" OR "conversational assistant*" OR "digital assist*" OR "intelligent assist*" OR "virtual assist*" OR "smart assist*" OR "virtual health assist*" OR "AI advisor*" OR "virtual advisor*" OR "animated advisor*" OR "smart advisor*" OR "AI coach*" OR "virtual coach*" OR "smart coach*" OR "animated coach*" OR "artificial conversation entit*" OR "Assistance technolog*" OR "conversational AI" OR "conversational interface*" OR "conversational system*" OR "dialogue system*" OR "natural language interface*" OR "automated conversation" OR "virtual conversation" OR ChatGPT OR Llama OR DeepSeek ) AND LA english NOT PT ( systematic review or meta analysis or meta-analysis or scoping review, or review ) |
| **Number** | **Original search scope: Physical activity, sleep, and sedentary**  Records identified in the original search: N = 201 (6/18/2025)  **After protocol revision: Physical activity only**  Updated total records identified: N = 206 (1/27/2026)  Newly added records after duplicate removal: N = 7 (1/27/2026) |
| **Note** | Search by Abstract and Title (XB) |

***IEEE Xplore (.ris)***

| **Search date** | 1/27/2026 |
| --- | --- |
| **Search terms** | lifestyle* OR "health behavior*" OR exercise* OR "physical activ*" |
|  | AND (Chatbot* OR chat-bot OR chat-bots OR "chat bot" OR "chat bots" OR "chat robot" OR"social bot" OR "social bots" OR eCoach OR infobot OR infobots OR "health bot" OR "health bots" OR smartbot OR smartbots OR "conversational bot" OR "conversational bots" OR "artificial intelligence chatbot" OR "artificial intelligence chatbots" OR "Ai agent" OR "Ai agents" OR "conversational agent" OR "conversational agents" OR "dialogue agent" OR "dialogue agents" OR "dialog agent" OR "dialog agents" OR "interactive agent" OR "interactive agents" OR "virtual agent" OR "virtual agents" OR "automated agent" OR "automated agents" OR "relational agent" OR "relational agents" OR "AI assistance" OR "AI assistant" OR "AI assistants" OR "conversational assistant" OR "conversational assistants" OR "digital assist" OR "digital assistant" OR "digital assistants" OR "digital assistance" OR "intelligent assist" OR "intelligent assistant" OR "intelligent assistants" OR "intelligent assistance" OR "virtual assist" OR "virtual assistance" OR "virtual assistant" OR "virtual assistants" OR "smart assist" OR "smart assistance" OR "smart assistant" OR "smart assistants" OR "dialogue agent" OR "dialogue agents" OR "dialog agent" OR "dialog agents" OR "AI advisor" OR "AI advisors" OR "virtual advisor" OR "virtual advisors" OR "smart advisor" OR "smart advisors" OR "AI avatar" OR "AI avatars" OR "virtual avatar" OR "virtual avatars" OR "animated avatar" OR "animated avatars" OR "smart avatar" OR "smart avatars" OR "AI coach" OR "AI coaches" OR "virtual coach" OR "virtual coaches" OR "smart coach" OR "smart coaches" OR "Assistance technologies" OR "conversational AI" OR "conversational interface" OR "conversational interfaces" OR "conversational system" OR "conversational systems" OR "Dialog system" OR "Dialog systems" OR "dialogue system" OR "dialogue systems" OR "natural language interface" OR "natural language interfaces" OR "automated conversation" OR "automated conversations" OR "virtual conversation" OR "virtual conversations" OR ChatGPT OR Llama OR DeepSeek) |
| **Filtering** | break down by “Conferences” and “Journals” |
| **Numbers** | **Original search scope: Physical activity, sleep, and sedentary**   - Records identified in the original search: N = 281 (6/18/2025)   **After protocol revision: Physical activity only**   - Updated total records identified: N = 274 (1/27/2026) - Newly added records after duplicate removal: N = 27 (1/27/2026) |
| **Note** | Search by abstract |

***ACM Digital Library (.enw)***

| **Search date** | 1/27/2026 |
| --- | --- |
| **Search terms** | lifestyle* OR "health behavior*" OR exercise* OR "physical activ*" |
|  | Chatbot* OR chat-bot* OR "chat bot*" OR "social bot*" OR eCoach* OR infobot* OR smartbot* OR "conversational bot*" OR "artificial intelligence chatbot*" OR "AI agent*" OR "conversational agent*" OR "dialogue agent*"OR "dialog agent*" OR "artificial agent" OR "interactive agent*" OR "virtual agent*" OR "automated agent*" OR "relational agent*" OR "dialogue agent*" OR "dialog agent*" OR "AI assist*" OR "virtual assist*" OR "conversational assistant*" OR "digital assist*" OR "intelligent assist*" OR "virtual assist*" OR "smart assist*" OR "virtual health assist*" OR "AI advisor*" OR "virtual advisor*" OR "animated advisor*" OR "smart advisor*" OR "AI coach*" OR "virtual coach*" OR "smart coach*" OR "animated coach*" OR "artificial conversation entit*" OR "Assistance technolog*" OR "conversational AI" OR "conversational interface*" OR "conversational system*" OR "dialogue system*" OR "natural language interface*" OR "automated conversation" OR "virtual conversation" OR ChatGPT OR Llama OR DeepSeek |
|  | break down by “Research Article” and “Short paper” |
| **Numbers** | **Original search scope: Physical activity, sleep, and sedentary**   - Records identified in the original search: N = 176 (6/18/2025)   **After protocol revision: Physical activity only**   - Updated total records identified: N = 253 (1/27/2026) - Newly added records after duplicate removal: N = 172 (1/27/2026) |
| **Note** | Search by abstract |

***ArXiv***

| **Search date** | 1/27/2026 |
| --- | --- |
| **Search terms** | ((lifestyle OR "health behavior" OR exercise OR "physical activity") AND (Chatbot OR "chat bot" OR "social bot" OR eCoach* OR infobot OR smartbot OR "conversational bot" OR "artificial intelligence chatbot" OR "AI agent" OR "conversational agent" OR "dialogue agent" OR "dialog agent" OR "artificial agent" OR "interactive agent" OR "virtual agent" OR "automated agent" OR "relational agent" OR "AI assist" OR "virtual assist" OR "conversational assistant" OR "digital assist" OR "intelligent assist" OR "smart assist" OR "virtual health assist" OR "AI advisor" OR "virtual advisor" OR "animated advisor" OR "smart advisor" OR "AI coach" OR "virtual coach" OR "smart coach" OR "animated coach" OR "artificial conversation entity" OR "Assistance technology" OR "conversational AI" OR "conversational interface" OR "conversational system" OR "dialogue system" OR "natural language interface" OR "automated conversation" OR "virtual conversation" OR ChatGPT OR Llama OR DeepSeek)) |
| **Numbers** | **Original search scope: Physical activity, sleep, and sedentary**   - Records identified in the original search: N = 178 (6/18/2025)   **After protocol revision: Physical activity only**   - Updated total records identified: N = 171 (1/27/2026) - Newly added records after duplicate removal: N = 42 (1/27/2026) |
| **Note** | Search by abstract |

***Med-arxiv***

| **Search date** | 1/27/2026 |
| --- | --- |
| **Search terms** | "lifestyle AND (Chatbot OR "conversational agent")"  "exercise AND (Chatbot OR "conversational agent")"  "health behavior AND (Chatbot OR "conversational agent")"  "physical activity AND (Chatbot OR "conversational agent")" |
| **Numbers** | **Original search scope: Physical activity, sleep, and sedentary**   - Records identified in the original search: N = 374 (6/18/2025)   **After protocol revision: Physical activity only**   - Updated total records identified: N = 455 (1/27/2026) - Newly added records after duplicate removal: N = 126 (1/27/2026) |
| **Note** | Search by title and abstract |

**Table S2.** Study eligibility criteria

| **Category** | **Inclusion criteria** | **Exclusion criteria** |
| --- | --- | --- |
| ***Study characteristics*** | | |
| P (Population) | Any gender, age, and ethnicity, from any country | None |
| I (Intervention) | A chatbot/conversational agent developed using existing AI or non-AI algorithms, models, or platforms. | Interventions that were not chatbot- or conversational agent-based, such as robotic interventions, wearable activity trackers, ecological momentary assessment, and message-based interventions. |
| C (Comparison) | No specific control group required | None |
| O (Outcomes) | Physical activity | Other health related behaviors, such as sedentary, sleep, diet, etc. |
| S (Study types) | Randomized controlled trail and non-randomized intervention studies, with or without a control group. | Review papers (e.g., relist review, systematic review, scoping review, meta-analysis, mixed methods review, concept analyses), observational study (e.g., cross-sectional, cohort, case-control), qualitative study, descriptive study (e.g., case reports), editorials, opinion pieces, conference abstracts, dissertations |
| ***Publication characteristics*** | | |
| Publication dates  Language  Publication status | Before 1/27/2026 | None |
|  | English | Other Language |
|  | Full online and Preprint | None |

**Table S3.** Data Extraction Table

| **Publication** | **Description** |
| --- | --- |
| Author (Year) | First author (online year) |
| Country | Corresponding author’s country |
| Study type | Different study designs, such as parallel randomized controlled trials, cluster randomized controlled trials, and pre-post studies. |
| **Exposure and Outcome** |  |
| Physical activity | (1) daily steps; (2) moderate-to-vigorous physical activity (MVPA, minutes/week); (3) total physical activity minutes (any measure of low, moderate and/or vigorous physical activity reported as a duration, e.g., minutes per day or week), and other physical activity related outcomes if available. |
| Measurement of outcome | The measurement method used to assess physical activity, such as self-report questionnaires or wearable activity trackers. |
| Chatbot algorithm | The algorithm used by the chatbot, such as non-AI algorithms, traditional AI algorithms, or large language models. |
| Chatbot output type | The chatbot delivery modality, such as text-based, voice/video-based, or mixed-modality chatbots. |
| Platform | The platform through which the chatbot interacts with users, such as a mobile phone, desktop computer, laptop, or tablet. |
| Chatbot visual expression | Chatbots with visual expressions, such as embodied conversational agents, and non-visual chatbots that interact through text or voice only. |
| **Population** |  |
| Target population | Report the target population and whether it is clinical or non-clinical, such as individuals with overweight or obesity, individuals with mental disorders, primary-school students, undergraduate students, working adults, older adults, people with disabilities, and disadvantaged populations. |
| #Participation | The total number of recruited participants. |
| %female | The percentage of female participants. |
| Age | The average age of recruited participants. |
| **Intervention** |  |
| Multicomponent | Whether the intervention was multicomponent, such as whether the chatbot was delivered alone or in combination with human support. |
| Brief description of the intervention | Description of the treatment provided in each study arm. |
| Intervention duration | The duration of the intervention. |
| Intervention setting | The setting in which the intervention was delivered, such as free-living settings, laboratory settings, or community spaces. |
| Chatbot interaction | How often participants interacted with the chatbot and how long each interaction lasted. |
| **Data extracted for the meta-analysis** | |
| #Participation | The sample size of the intervention and control groups at each specific time point. |
| Mean | The mean value of a specific outcome at a specific time point. |
| Standard deviation | The standard deviation of a specific outcome at a specific time point. |
| Assessment time | The timing of outcome assessment, such as mid-intervention, post-intervention, or follow-up, including the specific time point. |

**Table S4.** Descriptions of the Study Arms

| **Author year** | **Outcomes (Measurement)** | **Frequency** | **Duration** | **Narm** | **Common** | **Arm0** | **Arm1** | **Arm2** | **Arm3** |
| --- | --- | --- | --- | --- | --- | --- | --- | --- | --- |
| Bickmore 2005A^1^ | Steps (pedometer) | daily | 5–10 minutes | 2 | Standard behavior interventions: 1. pedometer; 2. log sheet for pedometers (manual log); 3. progress charts showing their activity levels over time; 4. daily web pages of educational content on the topic of walking for exercise | Standard behavior interventions | Relational ECA intervention group: set daily steps goals and help overcome barriers to PA | N.A. | N.A. |
| Bickmore 2005B^2^ | MVPA (self-report); Steps (self-report pedometer) | daily | 5–10 minutes | 3 | Standard behavior interventions: 1. pedometer; 2. log sheet for pedometers (manual log); 3. progress charts showing their activity levels over time; 4. daily web pages of educational content on the topic of walking for exercise | Standard behavior interventions | Non-relational ECA | Relational ECA | N.A. |
| Bickmore 2010^3^ | Steps (pedometer) | daily | 5–10 minutes | 2 | 1. pedometer automatically syncs to the system; 2. daily web pages of educational content on the topic of walking for exercise | Relational ECA (Non-variability: dialogue type and agent appearance do not change) | Relational ECA (Variability) | N.A. | N.A. |
| Bickmore 2013A^4^ | Steps (pedometer) | daily | 5–10 minutes | 2 | Standard behavior interventions: 1. pedometers; 2. daily web pages of educational content on the topic of walking for exercise | Standard behavior interventions | Relational ECA | N.A. | N.A. |
| Bickmore 2013B^5^ | Steps (pedometer) | daily | 5–10 minutes | 4 | pedometer | Pedometer only | Relational ECA targeting on exercise only | Relational ECA targeting diet only | Relational ECA targeting exercise and diet together |
| King 2013^6^ | Walking minutes (self-report) | weekly | 5–10 minutes | 2 | pedometer | General health promotion program at the community center | Relational ECA | N.A. | N.A. |
| King 2020^7^ | Steps (pedometer); Walking minutes, MVPA, Total PA time in minutes (self-report survey) | 2 times per month | 10-15 minutes | 2 | 1. pedometer; 2. introductory session includes national guidelines and safety tips | Human advisor | Relational ECA | N.A. | N.A. |
| Watson 2012^8^ | Steps (pedometer) | 3 times per week | 5-10 minutes | 2 | Standard behavior interventions: 1. pedometer; 2. website | Standard behavior interventions | Relational ECA | N.A. | N.A. |
| Op Den Akker 2016^9^ | PA levels (activity sensor) | daily | N.R. | 3 | Activity sensor | Web-based: receive text feedback inactively through website | Text chatbot: receive text feedback activity through coaching application | Relational ECA: receive feedback actively through ECA | N.A. |
| Friederichs 2014^10^ | MVPA (self-report) | one-time | N.R. | 3 | No | No intervention | Text chatbot: no avatar | Relational ECA | N.A. |
| Chauvin 2023^11^ | Total PA in MET-minutes (self-report) | one-time | N.R. | 3 | No | Text-based website | Relational ECA (empathetic) | Relational ECA (positive reframing) | N.A. |
| Aydemir 2025^12^ | Total PA time, including light PA and MVPA (self-report) | N.R. | N.R. | 2 | No | No intervention | ChatGPT | N.A. | N.A. |
| Carlin 2021^13^ | Total PA time, including light PA and MVPA (Actigraph GT3); Exercise duration (self-report) | daily | N.R. | 2 | Community-based weight management program (SWEET) | Community-based weight management program | Amazon Alexa | N.A. | N.A. |
| Chopra 2024^14^ | Exercise duration (N.R.) | weekly | N.R. | 2 | No | No intervention | DigiMate Customized Coach using ChatGPT API. | N.A. | N.A. |
| Collombon 2024^15^ | MVPA (Actigraph GT3X-BT; self-report) | 3 prompts/day in Week 1, decreasing to 1 prompt/day in the final week. | N.R. | 7 | AP: computer-tailored intervention-based theory of planned behavior, self-regulation theory, precaution adoption process model; IM: computer-tailored intervention based on self-determination theory and motivational interviewing | No intervention | AP/IM + Xiaomi band | AP/IM + Ecological momentary assessment | AP/IM + chatbot feedback |
| Hassoon 2021^16^ | Steps (Fitbit charge 2 HR) | 2 times per day | N.R. | 3 | Fitbit Charger | Education materials | Audio chatbot (Amazon Echo/ Alexa smart speaker) | Text chatbot | N.A. |
| Kohlbrenner 2024^17^ | Steps (Fitbit charge 3) | daily | 15 min/interaction for the intervention group; 13 min/interaction for the control group. | 2 | Standard behavior interventions: 1. CAir Desk for home-monitoring, 2. usual care (ice, physician visits, prescribed medication) | Standard behavior interventions | Receive additional physical activity feedback from chatbot | N.A. | N.A. |
| Kramer 2020^18^ | Steps (Smartphone build-in tracker); % of total step-goal achievement (Smartphone build-in tracker) | weekly | N.R. | 3 | Incentives and self-monitoring | Chatbot without planning feature | Chatbot with coping planning | Chatbot with action planning | N.A. |
| Jörke 2025^19^ | Total PA in MET-minutes (self-report) | N.A. | N.A. | 2 | 1. Apple Kit; 2. other functions on the Bloom App (i.e., ambient display, push notifications) | Bloom App without LLM control | Bloom with LLM control | N.A. | N.A. |
| Persell 2020^20^ | MVPA (self-report) | N.R. | N.R. | 2 | Home blood pressure monitoring (HBPM) | HBPM | Omron chatbot coaching App | N.A. | N.A. |
| Peuters 2024^21^ | Euclidean Norm Minus One (Axivity AX3) | any | any | 2 | No | No intervention | 1. Chatbot coaching App; 2. Other functions in the APP; 3. Fitbit | N.A. | N.A. |
| Rodrigues 2024^22^ | Steps (Apple HeathKit and Google Fit) | daily | N.R. | 4 | 1. smartphone build-in activity tracker | Chatbot without human cues | Chatbot with visual cues only | Chatbot with relational cues | Chatbot with visual and relational cues |
| Sun 2025^23^ | Steps (Activity tracking via App) | daily | 5 minutes | 3 | 1. other app functions such as activity tracking interface | Rule-based chatbot + LLM (prompt) chatbot | Rule-based chatbot + LLM (retrieval-augmented generation) chatbot | N.A. | N.A. |
| Sun 2024^24^ | Steps (Smartphone build-in tracker) | 1 time per 2 days | N.R. | 3 | no | No intervention | Chatbot (non-humorous) | Chatbot (humorous) |  |
| Dan 2026^25^ | Total PA in MET-minutes and total activity minutes (self-report; Actigraph GT3X) | any | N.R. | 2 | no | ChatGPT (protocol prompt) | ChatGPT (personalized prompt) | N.A. | N.A. |
| Oh 2025^26^ | Steps (Smartphone build-in tracker) | daily | N.R. | 2 | Daily steps report | Chatbot (educational sessions + non-relational cues) | Chatbot (educational sessions + relational cues) | N.A. | N.A. |
| Zheng 2026^27^ | MVPA (self-report) | 30 brief sessions in total | 5–8 minutes | 4 | Other app-based components | Base LvL UP app + other educational resources | LvL UP app with a peer supporter–LvL UP Buddy | N.A. | N.A. |
| Abdulrahman 2019^28^ | Intention to change PA (self-report) | one-time | 10 minutes | 3 | No | No intervention | ECA (unexplainable) | ECA (explainable) | N.A. |
| Abdulrahman 2022^29^ | Intention to change PA (self-report) | one-time | 10 minutes | 3 | No | ECA (belief-based explanation) | ECA (goal-based explanation) | ECA (belief- and goal-based explanation) | N.A. |
| Olafsson 2019^30^ | Self-efficacy in PA (self-report) | one-time | NR | 2 | No | ECA (coerced → not coerced) | ECA (not coerced → coerced) | N.A. | N.A. |
| Albers 2023^31^ | Self-efficacy in PA (self-report) | one-time | 7 minutes | 2 | No | Chatbot (generic dialogue) | Chatbot (personalized dialogue) | N.A. | N.A. |
| Galland 2025^32^ | Decisional balance toward PA (one item self-report) | one-time | 10 minutes | 2 | No | ECA (LLM only without external dialog management) | ECA (LLM with external dialog management) | N.A. | N.A. |
| Gardiner 2017^33^ | PA as a stress management technique (one item self-report) | daily | any duration | 2 | No | Human recordings | ECA conversation | N.A. | N.A. |
| Piao 2020^34^ | Self-Report Habit Index on PA (self-report) | daily | any duration | 2 | No | Chatbot (half intrinsic rewards and full extrinsic rewards) | Chatbot (half intrinsic and extrinsic rewards) | N.A. | N.A. |
| Caru 2023^35^ | Steps (pedometer) | daily | <5 minutes | 1 | The chatbot performs as delivery platform. All participants will receive interventions, such as CBT, mindfulness, exercise videos. | N.A. | Amazon Alexa | N.A. | N.A. |
| Chew 2024^36^ | Total PA in MET-minutes (self-report) | 3 times per day | N.R. | 1 | 1. food-based computer vision image recognition; 2. automated time-based nudges and meal stopwatch | N.A. | EtripApp with chatbot-based check in | N.A. | N.A. |
| Dhinagaran 2021^37^ | Total PA in MET-minutes (self-report) | Four times per day, once for each topic. | N.R. | 1 | No | N.A. | Chatbot (provide advice) | N.A. | N.A. |
| Davis 2020^38^ | MVPA (self-report) | weekly | N.R. | 1 | 1. induction session; 2. purpose-designed website; 3. activity tracker with weekly log sheet | N.A. | Chatbot (1. teach knowledge; 2. guide weekly check-in; 3. 24/7 on-time support) | N.A. | N.A. |
| Maher 2020^39^ | MVPA (self-report) | weekly | N.R. | 1 | 1. induction session; 2. purpose-designed website; 3. activity tracker with weekly log sheet | N.A. | Chatbot (1. teach knowledge; 2. guide weekly check-in; 3. 24/7 on-time support) | N.A. | N.A. |
| To 2021^40^ | Steps (Fitbit Flex 1); Total PA time in minutes (self-report) | 2–3 messages per day | N.R. | 1 | Fitbit self-monitoring | N.A. | Chatbot (1. provide PA feedback, 2. send motivational messages 3. help adjust PA goals; 4. provide educational resources; 5. track PA history) | N.A. | N.A. |
| Joseph 2023^41^ | Total PA in hours per day (self-report) | N.R. | N.R. | 1 | No | N.A. | Chatbot (1. boost participant health-literacy levels; 2. encourage the formation/continuation of health promoting behaviors) | N.A. | N.A. |
| Tan 2026^42^ | Exercise duration (self-report) | 3 times per week | N.R. | 1 | Proudme Tech intervention | N.A. | Chatbot (provide real-time context-aware feedback) | N.A. | N.A. |
| Mauricio 2025^43^ | Steps (Smartphone build-in tracker) | at least 2 times per day | N.R. | 1 | Other app functions | N.A. | Chatbot (provide recommendations) | N.A. | N.A. |

**Table S5.** Subgroup analysis

| **Outcome** | **Subgroup** | **_level** | **n_study** | **n_nodes** | **Hedges_g** | **CI_low** | **CI_high** | **p_value** | **subgroup_df** | **subgroup_p** |
| --- | --- | --- | --- | --- | --- | --- | --- | --- | --- | --- |
| All | Multiple intervention | No | 3 | 4 | 0.47 | -0.27 | 1.21 | 0.216 | 1 | 0.665 |
| All | Multiple intervention | Yes | 21 | 44 | 0.30 | 0.18 | 0.43 | 0.000 | 1 | - |
| All | Visual expression | No | 18 | 34 | 0.28 | 0.14 | 0.42 | 0.000 | 1 | 0.516 |
| All | Visual expression | Yes | 6 | 14 | 0.38 | 0.13 | 0.62 | 0.003 | 1 | - |
| All | Output type | Text | 17 | 29 | 0.28 | 0.13 | 0.42 | 0.000 | 2 | 0.690 |
| All | Output type | Audio | 3 | 6 | 0.25 | -0.26 | 0.77 | 0.335 | 2 | - |
| All | Output type | Mix | 5 | 13 | 0.40 | 0.15 | 0.66 | 0.002 | 2 | - |
| All | Algorithm type | Non-AI | 11 | 27 | 0.24 | 0.09 | 0.39 | 0.002 | 2 | 0.253 |
| All | Algorithm type | Traditional AI | 10 | 18 | 0.44 | 0.26 | 0.61 | 0.000 | 2 | - |
| All | Algorithm type | LLM | 3 | 3 | 0.42 | -0.68 | 1.51 | 0.455 | 2 | - |
| All | Comparison group | No intervention | 2 | 3 | 0.64 | -0.32 | 1.60 | 0.189 | 4 | **0.008** |
| All | Comparison group | Usual care | 8 | 17 | 0.29 | 0.12 | 0.46 | 0.001 | 4 | - |
| All | Comparison group | Digital non-chatbot | 3 | 4 | -0.06 | -0.22 | 0.10 | 0.468 | 4 | - |
| All | Comparison group | Text chatbot (base) | 3 | 8 | 0.03 | -0.11 | 0.16 | 0.705 | 4 | - |
| All | Comparison group | Human advisor | 1 | 4 | 0.61 | 0.01 | 1.21 | 0.045 | 4 | - |
| All | Duration | ≤6 weeks | 9 | 23 | 0.25 | 0.11 | 0.39 | 0.000 | 2 | 0.540 |
| All | Duration | 6–12 weeks | 9 | 14 | 0.36 | 0.03 | 0.69 | 0.032 | 2 | - |
| All | Duration | >12 weeks | 6 | 11 | 0.42 | 0.13 | 0.71 | 0.005 | 2 | - |
| All | Setting | Community | 2 | 5 | 0.71 | 0.19 | 1.24 | 0.008 | 2 | 0.141 |
| All | Setting | Home | 8 | 16 | 0.20 | 0.09 | 0.32 | 0.001 | 2 | - |
| All | Setting | Free-living | 13 | 26 | 0.30 | 0.14 | 0.45 | 0.000 | 2 | - |
| All | Participants clinical status | No | 16 | 35 | 0.33 | 0.18 | 0.48 | 0.000 | 1 | 0.746 |
| All | Participants clinical status | Yes | 7 | 9 | 0.37 | 0.17 | 0.57 | 0.000 | 1 | - |
| Steps | Multiple intervention | No | 1 | 2 | 0.17 | -0.32 | 0.66 | 0.496 | 1 | 0.814 |
| Steps | Multiple intervention | Yes | 11 | 20 | 0.23 | 0.13 | 0.33 | 0.000 | 1 | - |
| Steps | Visual expression | No | 7 | 14 | 0.25 | 0.11 | 0.39 | 0.001 | 1 | 0.795 |
| Steps | Visual expression | Yes | 5 | 8 | 0.22 | 0.07 | 0.37 | 0.005 | 1 | - |
| Steps | Output type | Text | 7 | 13 | 0.20 | 0.06 | 0.33 | 0.004 | 2 | 0.395 |
| Steps | Output type | Audio | 2 | 2 | 0.53 | 0.06 | 1.00 | 0.026 | 2 | - |
| Steps | Output type | Mix | 4 | 7 | 0.23 | 0.08 | 0.39 | 0.003 | 2 | - |
| Steps | Algorithm type | Non-AI | 8 | 16 | 0.17 | 0.05 | 0.28 | 0.004 | 1 | 0.075 |
| Steps | Algorithm type | Traditional AI | 4 | 6 | 0.35 | 0.18 | 0.52 | 0.000 | 1 | - |
| Steps | Comparison group | No intervention | 1 | 2 | 0.17 | -0.32 | 0.66 | 0.496 | 3 | 0.131 |
| Steps | Comparison group | Usual care | 5 | 9 | 0.33 | 0.13 | 0.54 | 0.002 | 3 | - |
| Steps | Comparison group | Text chatbot (base) | 3 | 6 | 0.04 | -0.15 | 0.22 | 0.707 | 3 | - |
| Steps | Comparison group | Human advisor | 1 | 1 | 0.42 | -0.01 | 0.85 | 0.057 | 3 | - |
| Steps | Duration | ≤6 weeks | 6 | 14 | 0.19 | 0.08 | 0.31 | 0.001 | 2 | 0.209 |
| Steps | Duration | 6–12 weeks | 2 | 2 | 0.94 | 0.12 | 1.76 | 0.025 | 2 | - |
| Steps | Duration | >12 weeks | 4 | 6 | 0.20 | 0.03 | 0.36 | 0.018 | 2 | - |
| Steps | Setting | Community | 1 | 1 | 0.42 | -0.01 | 0.85 | 0.057 | 2 | 0.519 |
| Steps | Setting | Home | 6 | 9 | 0.27 | 0.10 | 0.44 | 0.002 | 2 | - |
| Steps | Setting | Free-living | 5 | 12 | 0.19 | 0.06 | 0.32 | 0.004 | 2 | - |
| Steps | Participants clinical status | No | 8 | 16 | 0.19 | 0.09 | 0.29 | 0.000 | 1 | **0.047** |
| Steps | Participants clinical status | Yes | 3 | 4 | 0.68 | 0.20 | 1.15 | 0.005 | 1 | - |
| MVPA | Visual expression | No | 5 | 7 | 0.14 | -0.04 | 0.32 | 0.128 | 1 | 0.123 |
| MVPA | Visual expression | Yes | 2 | 3 | -0.08 | -0.28 | 0.13 | 0.476 | 1 | - |
| MVPA | Output type | Text | 5 | 7 | 0.14 | -0.04 | 0.32 | 0.128 | 1 | - |
| MVPA | Output type | Mix | 2 | 3 | -0.08 | -0.28 | 0.13 | 0.476 | 1 | - |
| MVPA | Algorithm type | non-AI | 4 | 6 | -0.06 | -0.19 | 0.08 | 0.402 | 1 | **0.004** |
| MVPA | Algorithm type | Traditional AI | 3 | 4 | 0.34 | 0.10 | 0.58 | 0.005 | 1 | - |
| MVPA | Comparison group | Usual care | 2 | 3 | 0.16 | -0.03 | 0.36 | 0.098 | 2 | 0.134 |
| MVPA | Comparison group | Digital non-chatbot | 2 | 3 | -0.04 | -0.21 | 0.13 | 0.618 | 2 | - |
| MVPA | Comparison group | Human advisor | 1 | 1 | -0.13 | -0.38 | 0.12 | 0.302 | 2 | - |
| MVPA | Duration | ≤6 weeks | 1 | 2 | 0.04 | -0.32 | 0.40 | 0.816 | 2 | 0.845 |
| MVPA | Duration | 6–12 weeks | 4 | 6 | 0.15 | -0.10 | 0.39 | 0.238 | 2 | - |
| MVPA | Duration | >12 weeks | 2 | 2 | 0.04 | -0.29 | 0.38 | 0.799 | 2 | - |
| MVPA | Setting | Community | 1 | 1 | -0.13 | -0.38 | 0.12 | 0.302 | 2 | 0.155 |
| MVPA | Setting | Home | 2 | 3 | 0.16 | -0.03 | 0.36 | 0.098 | 2 | - |
| MVPA | Setting | Free-living | 4 | 6 | 0.15 | -0.10 | 0.39 | 0.238 | 2 | - |
| MVPA | Participants clinical status | No | 4 | 6 | 0.18 | -0.09 | 0.46 | 0.193 | 1 | 0.946 |
| MVPA | Participants clinical status | Yes | 2 | 2 | 0.17 | -0.03 | 0.37 | 0.098 | 1 | - |
| Total PA | Multiple intervention | No | 1 | 1 | -0.01 | -0.37 | 0.36 | 0.975 | 1 | 0.058 |
| Total PA | Multiple intervention | Yes | 5 | 7 | 0.52 | 0.12 | 0.93 | 0.012 | 1 | - |
| Total PA | Visual expression | No | 5 | 7 | 0.36 | -0.06 | 0.78 | 0.092 | 1 | **0.030** |
| Total PA | Visual expression | Yes | 1 | 1 | 0.90 | 0.64 | 1.17 | 0.000 | 1 | - |
| Total PA | Output type | Text | 4 | 5 | 0.41 | 0.00 | 0.82 | 0.050 | 2 | 0.125 |
| Total PA | Output type | Audio | 1 | 2 | 0.31 | -1.82 | 2.43 | 0.777 | 2 | - |
| Total PA | Output type | Mix | 1 | 1 | 0.90 | 0.64 | 1.17 | 0.000 | 2 | - |
| Total PA | Algorithm type | non-AI | 1 | 1 | 0.90 | 0.64 | 1.17 | 0.000 | 2 | **0.001** |
| Total PA | Algorithm type | AI | 4 | 6 | 0.47 | 0.05 | 0.89 | 0.030 | 2 | - |
| Total PA | Algorithm type | LLM | 1 | 1 | -0.23 | -0.76 | 0.31 | 0.405 | 2 | - |
| Total PA | Comparison group | Usual care | 1 | 2 | 0.31 | -1.82 | 2.43 | 0.777 | 2 | **0.001** |
| Total PA | Comparison group | Digital non-chatbot | 1 | 1 | -0.23 | -0.76 | 0.31 | 0.405 | 2 | - |
| Total PA | Comparison group | Human advisor | 1 | 1 | 0.90 | 0.64 | 1.17 | 0.000 | 2 | - |
| Total PA | Duration | ≤6 weeks | 4 | 5 | 0.41 | 0.00 | 0.82 | 0.050 | 2 | 0.125 |
| Total PA | Duration | 6–12 weeks | 1 | 2 | 0.31 | -1.82 | 2.43 | 0.777 | 2 | - |
| Total PA | Duration | >12 weeks | 1 | 1 | 0.90 | 0.64 | 1.17 | 0.000 | 2 | - |
| Total PA | Setting | Community | 1 | 1 | 0.90 | 0.64 | 1.17 | 0.000 | 2 | 0.125 |
| Total PA | Setting | Home | 1 | 2 | 0.31 | -1.82 | 2.43 | 0.777 | 2 | - |
| Total PA | Setting | Free-living | 4 | 5 | 0.41 | 0.00 | 0.82 | 0.050 | 2 | - |
| Total PA | Participants clinical status | No | 5 | 6 | 0.38 | -0.11 | 0.86 | 0.126 | 1 | 0.617 |
| Total PA | Participants clinical status | Yes | 2 | 2 | 0.61 | -0.17 | 1.40 | 0.126 | 1 | - |
| Exercise time | Multiple intervention | No | 1 | 1 | 1.57 | 1.00 | 2.13 | 0.000 | 1 | **0.000** |
| Exercise time | Multiple intervention | Yes | 2 | 3 | -0.05 | -0.26 | 0.15 | 0.608 | 1 | - |
| Exercise time | Output type | Text | 2 | 2 | 0.74 | -0.84 | 2.32 | 0.358 | 1 | 0.291 |
| Exercise time | Output type | Audio | 1 | 2 | -0.23 | -1.09 | 0.63 | 0.602 | 1 | - |
| Exercise time | Algorithm type | Traditional AI | 1 | 2 | -0.23 | -1.09 | 0.63 | 0.602 | 1 | 0.291 |
| Exercise time | Algorithm type | LLM | 2 | 2 | 0.74 | -0.84 | 2.32 | 0.358 | 1 | - |
| Exercise time | Comparison group | No intervention | 1 | 1 | 1.57 | 1.00 | 2.13 | 0.000 | 1 | **0.001** |
| Exercise time | Comparison group | Usual care | 1 | 2 | -0.23 | -1.09 | 0.63 | 0.602 | 1 | - |
| Exercise time | Setting | Home | 1 | 2 | -0.23 | -1.09 | 0.63 | 0.602 | 1 | **0.001** |
| Exercise time | Setting | Free-living | 1 | 1 | 1.57 | 1.00 | 2.13 | 0.000 | 1 | - |
| Exercise time | Participants clinical status | No | 3 | 3 | 0.51 | -0.60 | 1.62 | 0.365 | 1 | 0.299 |
| Exercise time | Participants clinical status | Yes | 1 | 1 | -0.35 | -1.55 | 0.84 | 0.564 | 1 | - |
| Walking | Comparison group | Usual care | 1 | 1 | 1.22 | 0.55 | 1.90 | 0.000 | 1 | 0.941 |
| Walking | Comparison group | Human advisor | 1 | 1 | 1.25 | 0.97 | 1.52 | 0.000 | 1 | - |

***Note.* n_study** = number of studies included in each analysis; **n_nodes** = number of effect sizes included in each analysis; **Hedges’ g** = standardized mean difference; **CI_low** = lower bound of the 95% confidence interval; **CI_high** = upper bound of the 95% confidence interval; **p_value** = p value for the pooled effect size; subgroup**_df** = degrees of freedom for the subgroup difference test; **subgroup_p** = p value for the subgroup difference test

**Table S6**. Meta-regression analysis (all PA outcomes)

| **Moderator** | **Category** | **n_study** | **n_nodes** | **beta** | **SE** | **CI_low** | **CI_high** | **p_value** | **QM** | **QM_p** |
| --- | --- | --- | --- | --- | --- | --- | --- | --- | --- | --- |
| Multiple intervention | Intercept (No) | 3 | 4 | 0.47 | 0.24 | 0.01 | 0.94 | 0.047 | 0.46 | 0.500 |
| Multiple intervention | Yes | 21 | 44 | -0.17 | 0.25 | -0.66 | 0.32 | 0.500 | **-** | **-** |
| Visual expression | Intercept (No) | 18 | 34 | 0.29 | 0.09 | 0.12 | 0.47 | 0.001 | 0.31 | 0.578 |
| Visual expression | Yes | 6 | 14 | 0.09 | 0.17 | -0.24 | 0.43 | 0.578 | **-** | **-** |
| Output type | Intercept (Text) | 17 | 29 | 0.28 | 0.10 | 0.10 | 0.47 | 0.003 | 0.67 | 0.716 |
| Output type | Audio | 3 | 6 | 0.02 | 0.28 | -0.53 | 0.56 | 0.956 | **-** | **-** |
| Output type | Mix | 5 | 13 | 0.15 | 0.18 | -0.21 | 0.50 | 0.416 | **-** | **-** |
| Algorithm type | Intercept (Non-AI) | 11 | 27 | 0.25 | 0.10 | 0.05 | 0.44 | 0.015 | 1.26 | 0.531 |
| Algorithm type | Traditional AI | 10 | 18 | 0.18 | 0.16 | -0.13 | 0.49 | 0.266 | **-** | **-** |
| Algorithm type | Generative AI (i.e., LLM) | 3 | 3 | 0.11 | 0.27 | -0.41 | 0.64 | 0.672 | **-** | **-** |
| Comparison group | Intercept (no intervention) | 2 | 3 | 0.81 | 0.32 | 0.18 | 1.43 | 0.012 | 6.24 | 0.100 |
| Comparison group | Usual care | 8 | 17 | -0.38 | 0.36 | -1.08 | 0.32 | 0.291 | **-** | **-** |
| Comparison group | Digital non-chatbot | 3 | 4 | -0.88 | 0.40 | -1.68 | -0.09 | 0.029 | **-** | **-** |
| Comparison group | Text chatbot (base) | 3 | 8 | -0.73 | 0.41 | -1.54 | 0.08 | 0.076 | **-** | **-** |
| Comparison group | Human advisor | 1 | 4 | **-** | **-** | **-** | **-** | **-** | **-** | **-** |
| Duration | Intercept (≤6 weeks) | 9 | 23 | 0.24 | 0.12 | 0.01 | 0.47 | 0.038 | 0.86 | 0.652 |
| Duration | 6–12 weeks | 9 | 14 | 0.13 | 0.18 | -0.23 | 0.49 | 0.480 | **-** | **-** |
| Duration | >12 weeks | 6 | 11 | 0.16 | 0.19 | -0.22 | 0.54 | 0.411 | **-** | **-** |
| Setting | Intercept (Community) | 2 | 5 | 0.74 | 0.22 | 0.31 | 1.17 | 0.001 | 3.96 | 0.138 |
| Setting | Home | 8 | 16 | -0.50 | 0.26 | -1.00 | 0.00 | 0.052 | **-** | **-** |
| Setting | Free-living | 13 | 26 | -0.43 | 0.24 | -0.90 | 0.04 | 0.072 | **-** | **-** |
| Setting | N.A. | 1 | 1 | **-** | **-** | **-** | **-** | **-** | **-** | **-** |
| Participants clinical status | Intercept (No) | 16 | 35 | 0.31 | 0.03 | 0.25 | 0.38 | 0.000 | 0.09 | 0.766 |
| Participants clinical status | Yes | 7 | 9 | 0.02 | 0.08 | -0.13 | 0.18 | 0.766 | **-** | **-** |
| Participants clinical status | N.A. | 2 | 4 | **-** | **-** | **-** | **-** | **-** | **-** | **-** |
| Assessment time | Intercept (Mid) | 4 | 7 | 0.18 | 0.15 | -0.10 | 0.47 | 0.208 | 1.92 | 0.382 |
| Assessment time | Post | 24 | 48 | 0.13 | 0.15 | -0.16 | 0.41 | 0.382 | - | - |
| Assessment time | Follow-up | 4 | 9 | -0.04 | 0.20 | -0.42 | 0.35 | 0.846 | - | - |
| Chatbot characteristics | NA | NA | NA | NA | NA | NA | NA | NA | 5.11 | 0.402 |
| Intervention characteristics | NA | NA | NA | NA | NA | NA | NA | NA | 9.31 | 0.749 |

***Note.* Chatbot characteristics** include output type, use of visual expression, and algorithm type; **Intervention characteristics** include intervention duration, intervention setting, and clinical status; **n_study** = number of studies included in each analysis; **n_nodes** = number of effect sizes included in each analysis; **beta** = estimated regression coefficient; **SE** = standard error; **CI_low** = lower bound of the 95% confidence interval; **CI_high** = upper bound of the 95% confidence interval; **p_value** = p value for the regression coefficient; **QM** = omnibus test statistic for moderators; **QM_p** = p value for the omnibus test.
